# The impacts of increased global vaccine sharing on the COVID-19 pandemic; a retrospective modelling study

**DOI:** 10.1101/2022.01.26.22269877

**Authors:** Sam Moore, Edward M. Hill, Louise Dyson, Michael J. Tildesley, Matt J. Keeling

## Abstract

**Background:** The SARS-CoV-2 pandemic has generated considerable morbidity and mortality world-wide. While the protection offered by vaccines (and booster doses) offers a method of mitigating the worst effects, by the end of 2021 the distribution of vaccine was highly heterogeneous with some countries achieving over 90% coverage in adults by the end of 2021, while others have less than 2%. In part, this is due to the availability of sufficient vaccine, although vaccine hesitancy also plays a role.

**Methods:** We use an age-structured model of SARS-CoV-2 dynamics, matched to national data from 152 countries, to investigate the global impact of different vaccine sharing protocols during 2021. We assume a direct relationship between the emergence of variants with increased transmissibility and the cumulative amount of global infection, such that lower global prevalence leads to a lower reproductive number within each country. We compare five vaccine sharing scenarios, from the current situation, through sharing once a particular within-country threshold is reached (e.g. all over 40s have received 2 doses), to full sharing where all countries achieve equal age-dependent vaccine deployment.

**Findings:** Compared to the observed distribution of vaccine uptake, we estimate full vaccine sharing would have generated a 1.5% (PI -0.1 - 4.5%) reduction in infections and a 11.3% (PI 0.6 - 23.2%) reduction in mortality globally by January 2022. The greatest benefit of vaccine sharing would have been experienced by low and middle income countries, who see an average 5.2% (PI 2.5% - 10.4%) infection reduction and 26.8% (PI 24.1% - 31.3%) mortality reduction. Many high income countries, that have had high vaccine uptake (most notably Canada, Chile, UK and USA), suffer increased infections and mortality under most of the sharing protocols investigated, assuming no other counter measures had been taken. However, if reductions in vaccine supply in these countries had been offset by prolonged use of non-pharmaceutical intervention measures, we predict far greater reductions in global infection and mortality of 64.5% (PI 62.6% - 65.4%) and 62.8% (PI 44.0% - 76.3%), respectively.

**Interpretation:** By itself, our results suggest that although more equitable vaccine distribution would have had limited impact on overall infection numbers, vaccine sharing would have substantially reduced global mortality by providing earlier protection of the most vulnerable. If increased vaccine sharing from high income nations had been combined with slower easing of non pharmaceutical interventions to compensate for this, a large reduction in both infection and mortality globally would be expected, confounded by a lower risk of new variants arising.

## Introduction

Since its emergence in Wuhan at the end of 2019, SARS-CoV-2 has rapidly spread around the world causing substantial epidemics in nearly every country. As the causative agent of COVID-19 disease, the virus has inflicted significant morbidity and mortality globally [1]. During 2020, the containment of the pandemic relied predominantly on non-pharmaceutical interventions (NPIs) to limit the spread of infection, reduce severe disease and help prevent health services from being overwhelmed [2, 3]. While this has broadly been effective, it has also been economically and socially damaging [4]. During late 2020 and early 2021 a number of vaccines were approved for public use, representing unparalleled development times; this has enabled many countries to implement mass vaccination campaigns as a means of mitigation.

To date (January 2022) approximately 49% of the global population have received a full dose of COVID vaccine, though delivery has varied greatly between (and within) countries [5, 6]. Many high-income countries have enjoyed very successful vaccination campaigns, with several exceeding 90% coverage of adults (aged 16 years and above). However, amongst many low and lower-middle income countries vaccine availability has been significantly more limited, with low income countries counting for as little as 0.9% of the overall total vaccine deployed [5].

Low and lower-middle income countries are mostly dependent on donations from wealthier nations and vaccine sharing schemes, such as the WHO directed COVAX initiative [7]. As an increasing number of countries begin to achieve high levels of vaccine coverage they face the decision of whether to continue with a nationalistic approach to vaccination — by extending rollout to the young, providing booster jabs to protect against waning immunity, and stockpiling surplus resources for future use — or whether to begin donating more vaccines to where there may be significantly higher pay-offs per dose in reducing infection and mortality. While high income nations typically have more elderly populations and consequently more individuals who are directly vulnerable to the effects of COVID-19, low income nations are typically less well equipped to deal with high levels of infection. The effects of increased pressure on already limited healthcare resources in poorer nations has had critical impacts on a range of endemic diseases [8, 9], and without surplus welfare resources available in such nations non-pharmaceutical interventions are unsustainable [10].

In the high income countries that have achieved high levels of vaccination, campaigns have proven highly effective in limiting disease impacts whilst allowing the relaxation of restrictions and a return to pre-pandemic-like behaviour [11, 12, 13]. However, the estimated waning of vaccine efficacy [14, 15, 16] has meant that many nations are now investing in booster campaigns [17] and considering fourth doses. The huge numbers of global infections (estimated to have affected 40-50% of the global population to date [18]) have generated considerable opportunities for viral mutation and the emergence of variants that have notable transmission and/or immune escape advantages over resident strains [19, 20]. Such variants of concern have prolonged the pandemic and often generated new waves of infection, raising the basic reproductive number and hence raising the herd immunity threshold. The continued threat of further mutation equates to a large level of uncertainty in future infection patterns. Consequently, while national vaccination campaigns have proven effective in limiting disease impact nationally, epidemic containment may only be fully achieved if high levels of new global infections are avoided, minimising the threat of generating further variants of concern [21, 22].

Through the use of our detailed global model, incorporating country level COVID-19 disease and SARS-CoV-2 vaccination data to the end of 2021 in 152 different countries, we explore the effects of increasing historic levels of global vaccine sharing on the likely state of the pandemic, projected from early 2020 to the end of 2021. We show that increased vaccine sharing may dramatically decrease mortality in lower income countries with only limited increases in some high income donor nations, provided the most vulnerable are still vaccinated in a timely manner. We also show the potential of more equally distributed vaccination; in return for an increased duration of control measures in the currently vaccine-rich minority of countries, vaccine sharing can substantially decrease the number of overall infections in 2021, reducing the potential for the evolution and spread of increasingly severe variants and subsequently dramatically improving the outlook of the pandemic globally.

## Methods and assumptions

We developed a mathematical model of SARS-CoV-2 transmission and COVID-19 disease outcomes, applied to 152 different countries, each with its own parameters reflecting demographics and social structure. The countries are simulated using an age-structured compartmental infection model – coupled by the global evolution of new variants and the sharing of vaccines when national conditions are met. Given the strong correlation between per capita income and the level of COVID-19 vaccination [23], we partition the 152 countries simulated into the four income group classifications given by the World Bank [24]. More details of the model structure and classification of countries may be found in the Supplementary Information.

Age is recognised to play an important role in the dynamics of the SARS-CoV-2 epidemic, strongly influencing both the outcome following infection and the characteristic social mixing behaviour [25]) that facilitates transmission. These age-based heterogeneities are captured by stratifying the modelled populations into 5 year age groups using country level data on age demographics [26], each with their own parameters for susceptibility, the occurrence of symptoms and risk of severe disease.

Throughout the pandemic, most countries have responded to rising levels of disease with mitigation measures including social distancing, quarantining, mandatory mask wearing, and contact tracing [27], while increased caution amongst residents may act to significantly slow viral spread [28]. To allow for these effects, epidemiologically relevant contacts within each country are varied in a time varying manner by a country-specific control factor.

Parameters, for national control measures and death rates, are fitted using daily estimates for infections and deaths proposed by the Institute for Health Metrics and Evaluation (IHME), together with levels of uncertainty [18]. Due to inconsistencies and under-reporting of COVID metrics in many countries, rather than relying on official reports, there estimates are made based on excess mortality statistics, comparing death rates in each country during the pandemic to historical data, tracking past trends and seasonality [29]. Other similar excess mortality estimates have been made elsewhere with reasonable consistency; for instance [30] estimates 160,000 deaths in South Africa by 27th June, 2021, (60,000 reported) compared to the IHME estimate of 156,373 deaths (with a high and low estimate range 90,221-258,352). Uncertainty in these values is accounted for by taking 100 independent random samples informed by the high and low IHME estimates, and propagating these samples through the fitting and simulation to generate means and 95% prediction intervals.

### Vaccination and sharing scenarios

We make the assumption that all countries aim to eventually achieve vaccine coverage in all individuals from the age of 12 and above, with a 90% uptake for those above the age of 60 and 80% for those below (although vaccine hesitancy may present substantial difficulty in achieving this in some nations). We also assume an oldest first approach to vaccine distribution is used in all nations, delivering vaccine to the most vulnerable first [31]. Although deviation from this approach has been seen in some nations that have chosen to prioritise essential workers or key disease spreaders (including those, like taxi drivers, in high contact professions), the strategy of oldest-first is the most widely employed [27].

Vaccination is assumed to provide individual protection against four measures: susceptibility, onward transmission, symptom probability and hospitalisation/death. These are based on efficacy characteristics similar to one or two doses of AstraZeneca (AZ) vaccine [14] (Fig. 1(a) and Table S1) – this being one of the most widely distributed and well studied COVID vaccines to date [32]. This is an approximation of the heterogeneous global picture [32], with some vaccines (such as Sinopharm) considered to provide lower protection [33] while the widely deployed Johnson & Johnson vaccine requires only a single dose [34].

**Figure 1:**
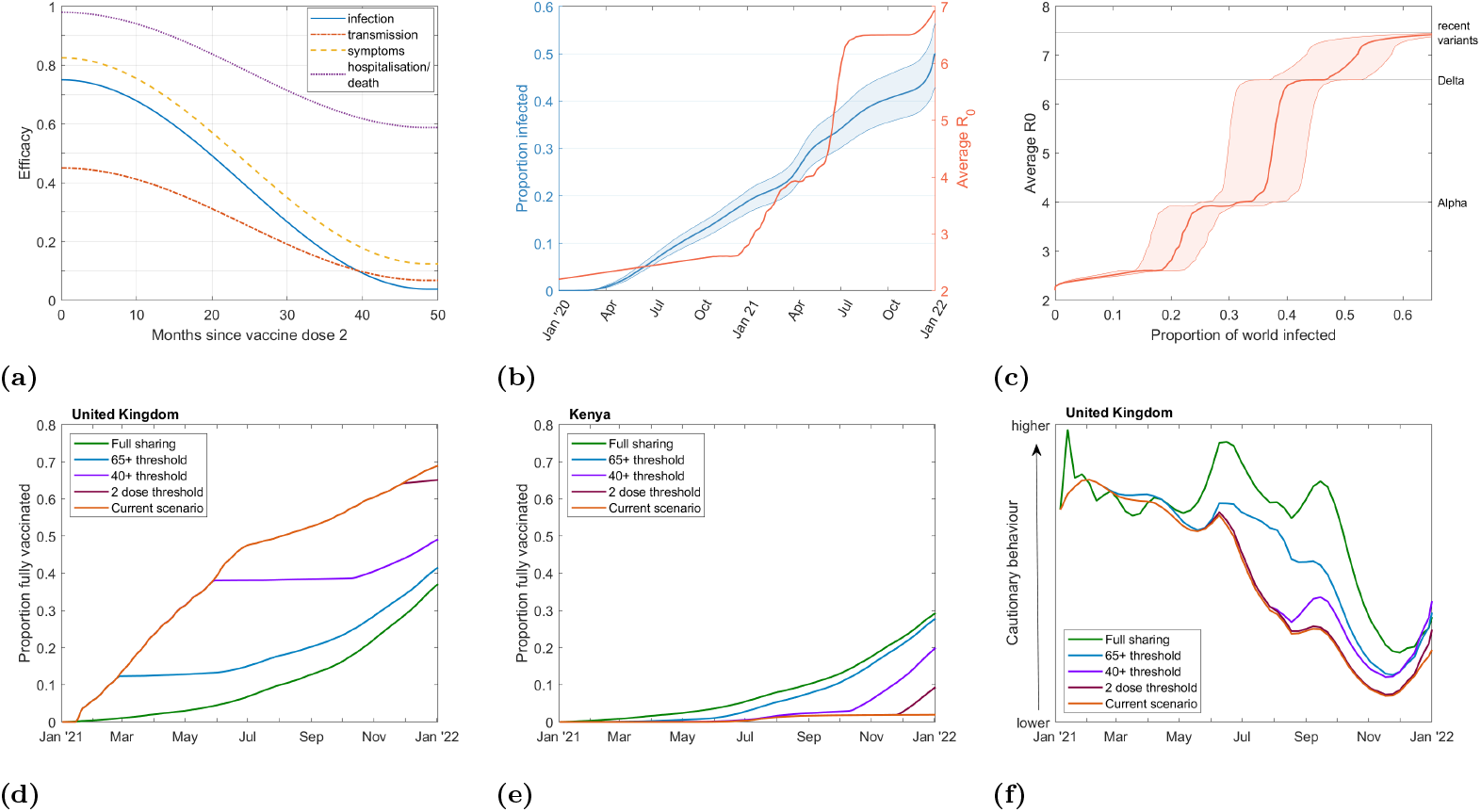
**(a)** Efficacy profiles used for vaccination following the second dose of vaccination. **(b, c)** Estimates for infected proportion of the global population (blue) and average *R*_0_ of SARS-CoV-2 strains in circulation (red). Higher and lower infection estimates are indicated by thinner outer lines with median estimates indicated by the thicker central line. Solid lines/shaded region are based on data to date, with the dotted lines/unshaded regions future projection. (b) shows separately the correlation between infection numbers with time (left axis, blue) and *R*_0_ with time (right axis, red), (c) shows the correlation between infected proportion and average *R*_0_. **(d, e)** Example plots comparing levels of vaccination over time in each scenario considered for a highly vaccinated nation (the United Kingdom) and a nation with limited vaccination (Kenya). **(f)** Example plot for a highly vaccinated nation (the United Kingdom) of levels of cautionary behaviour used for each level of vaccine sharing, for scenarios with adapted behaviour.

In addition, efficacy can also vary with age, dose interval and between variants. All four forms of vaccine protection are assume to wane over time, from a maximum shortly after the second dose to minimum levels 4 years (48 months) later, Fig. 1(a). Countries completing 2 dose vaccination coverage, and with sufficient vaccine supply, are assumed to commence delivery of booster vaccinations to all individuals six months after receiving their second dose, again in oldest to youngest priority order. Booster doses when delivered are taken to reset waning back to the maximum efficacy level. Alongside a default, low-sharing, scenario reflecting actual historical vaccine delivery, we additionally test a number of more collaborative strategies that begin at the start of 2021:

- **Current scenario, low sharing:** Past reported vaccination rates are followed in each country.
- **2-dose threshold:** The simulation progresses in each country, with vaccination rates equal to the *current scenario* until the 2-dose vaccination programme is completed. Subsequent vaccine deliveries from that country are then divided between all countries proportional to the number of unvaccinated individuals remaining in each.
- **40+ threshold:** Similar to the *2-dose threshold* scenario except vaccine sharing begins in each country after a 2-dose vaccination is completed for all individuals aged 40 and above.
- **65+ threshold:** Similar to the *2-dose threshold* scenario except vaccine sharing begins in each country after a 2-dose vaccination is completed for all individuals aged 65 and above.
- **Full sharing:** Vaccine sharing begins at the start of 2021 with all vaccination pooled and divided between all countries proportional to the number of unvaccinated individuals remaining in each.

### Adapted behaviour

In scenarios where some countries have reduced vaccine supply due to increased sharing, it is likely that, without a change to behaviour or controls, infections would increase compared to the current scenario. We therefore perform two projections, one where the behaviour follows inferred levels irrespective of infections and one where behaviour adapts. We implement this behavioural response by increasing/decreasing the control parameter for each country that is sharing vaccines, dependent on whether the number of active infections is increasing/decreasing (subject to a five day time lag to reflect delays in detection and reaction). An example showing behaviour adaptation is given in Fig. 1(f).

### Variants

Across the course of the pandemic there has been a substantial rise in the level of transmissibility of the dominant SARS-CoV-2 variant. Initially, when COVID-19 was first detected in China, the basic reproductive ratio, *R*_0_ was estimated between 2 and 2.4 [35]. Transmissibility has seen three major step changes due to: the Alpha variant (*R*_0_ ≈ 4 − 5) at the end of 2020; the Delta variant (*R*_0_ ≈ 6 − 7) [36, 37] becoming dominant in many countries by early summer 2021; and most recently the emergence of the Omicron variant.

By considering the global proportion of each variant (as averaged across countries for which such data are available from the GISAID database [38], assuming wild type has *R*_0_ = 2.2, Alpha variant has *R*_0_ = 4, and Delta has *R*_0_ = 6.5), we may visualise the trend of increasing *R*_0_ (Fig. 1(b)) and the associated level of infection up to that time. Variants recently succeeding Delta (including Omicron and Delta plus) are more difficult to quantify, and as such we take a conservative estimate of an associated increase for a further 15% transmission advantage associated with all such strains. The relationship between total historic infections (blue) and the average basic reproductive number (red), is then used to realise the impact of varying infection levels on variant emergence in the simulations – relating a given level of historic infection to an average basic reproductive ratio due to the emergence of new variants Fig. 1(c).

## Results

Nearly 50% of the global population have been fully vaccinated by the end of 2021, though with large disparities in coverage across the globe meaning this figure is closer to 75% across high income countries, but lower than 2% in many low income countries. Any increased degree of vaccine sharing will at least partially address this balance, potentially generating substantial gains in sparsely vaccinated nations, though inevitably leading to some loss in the most highly vaccinated countries (Fig. 2, Fig. 3).

**Figure 2:**
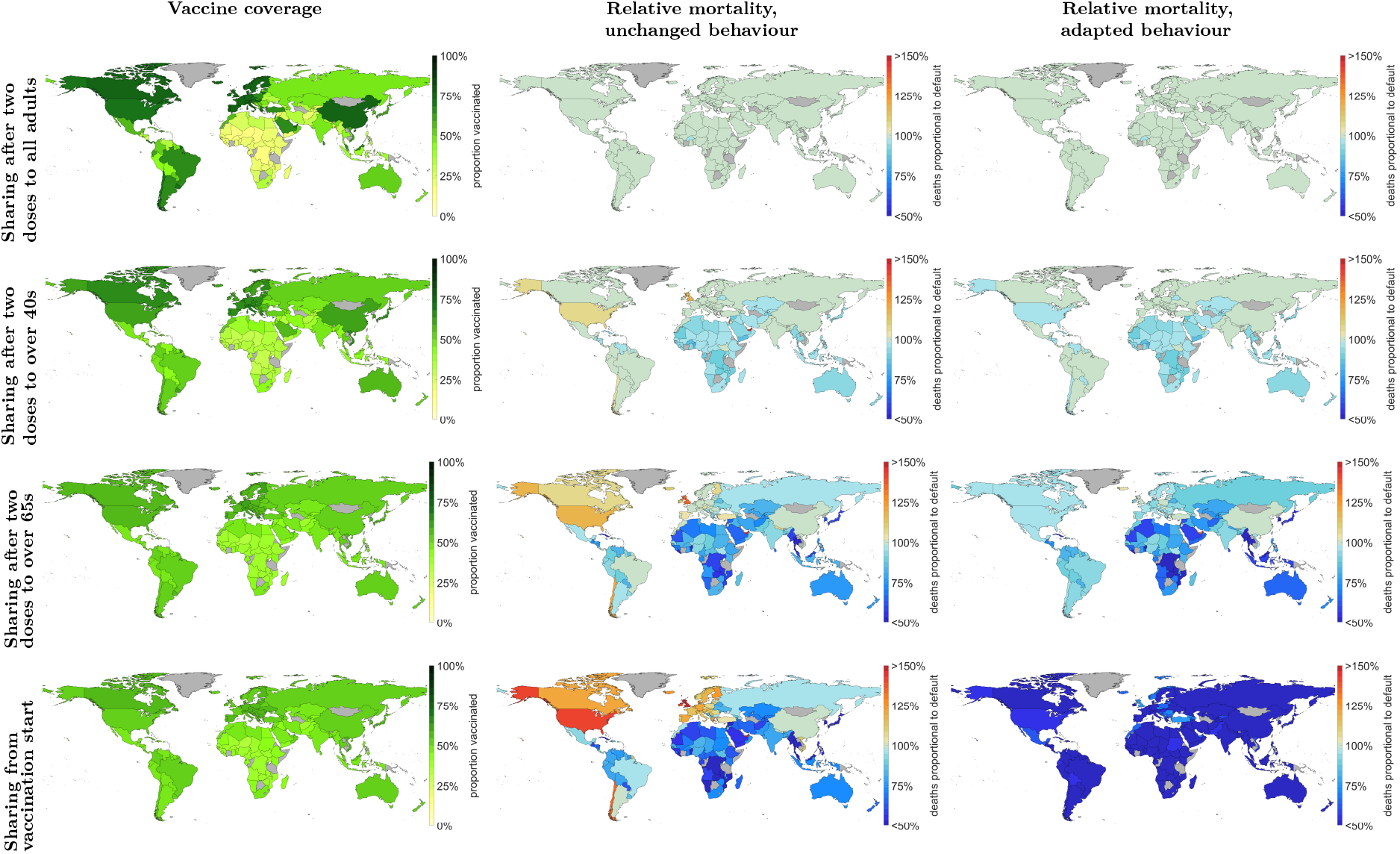
Country level estimates of vaccination coverage (left column), total number of deaths relative to the current scenario with unchanged behaviour but increased vaccine sharing (central column), and total number of deaths relative to the current scenario with unchanged behaviour but increased vaccine sharing (right hand column). All estimates are given as mean simulation values for the start of 2022, with 95% prediction intervals provided in Table S2. Analogous figures for infection estimates are given in Figure S3.

**Figure 3:**
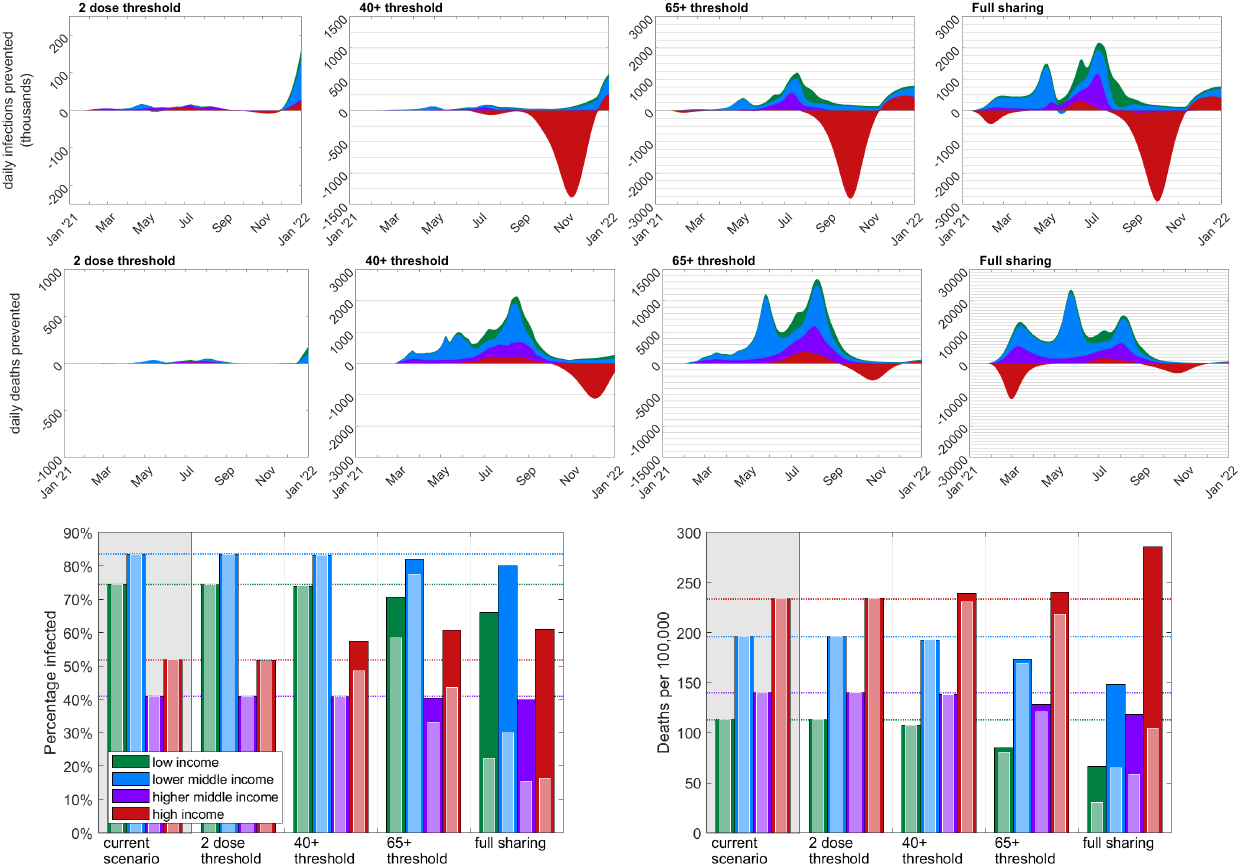
The top two rows give time series plots showing the reduction in the global number of daily infections and daily deaths compared to the default scenario. The different coloured areas represent the proportion of infections/deaths prevented in countries from each economic group: low income (green); lower middle income (blue), higher middle income (purple); high income (red). The columns show the four different vaccine sharing scenarios, each assuming unadapted behaviour. Equivalent figures for adapted behaviour are given in Fig S2. The lower plots show predicted total proportion infected (left) and deaths per 100,000 (right) from COVID-19 by the start of 2022 in each of the economic regions. The darker (rear) bars represent simulations where unadapted behaviour is followed, reflecting the default fitting in each. The lighter (front) bars represent simulations with adapted behaviour in countries commencing vaccine sharing. The first group of bars in each (grey shaded region) and dotted lines, correspond to the default scenario with subsequent bars representing simulations with the four different vaccine sharing strategies. All results presented here are simulation averages, with associated prediction intervals listed in Table 1.

If previous behaviour remains unchanged, we observe that increased vaccine sharing is likely to have led to benefits in low, lower middle and even higher middle income countries across early to mid-2021, working to reduce infections by 11.5%, 4.0% and 2.4% in these regions respectively in the full sharing scenario (Table S2). However, these benefits might have been largely offset by significant increases in infection experienced in high income countries later in 2021 as other control measures were relaxed (Fig. 3, top row), with approximately 17.2% more infections in the full sharing scenario. Australia and New Zealand appear as notable exceptions to this, as having relatively late starting vaccination programs means few doses are given away in any scenario, and very low infection levels to date mean that even small advantages from delayed variant emergence translate into large percentage gains (Fig. 2).

**Table 1:** Estimates for vaccination coverage, proportion infected and mortality rates (per 100,000) under each vaccine sharing strategy and income group taken at the start of 2022. Bracketed values represent a 95% prediction interval. Individual country estimates for all 152 simulated countries provided in the Supplementary Information.

| Region | Strategy | Proportion vaccinated<br>(2 doses) | Proportion infected |  | Mortalities per 100,000 |  |
| --- | --- | --- | --- | --- | --- | --- |
|  |  |  | unchanged<br>behaviour | adapted<br>behaviour | unchanged<br>behaviour | adapted<br>behaviour |
| World | Default | 43.9% | 60.5% (54.6%,63.9%) | 60.5% (54.6%,63.9%) | 189.8 (93.5,294.9) | 189.8 (93.5,294.9) |
|  | 2 dose threshold | 49.3% | 60.4% (54.5%,63.9%) | 60.4% (54.5%,63.9%) | 194.9 (106.0,294.7) | 194.9 (106.0,294.7) |
|  | 40+ threshold | 49.4% | 61.1% (55.2%,64.5%) | 59.8% (53.2%,63.6%) | 192.8 (105.8,290.7) | 191.6 (105.3,288.3) |
|  | 65+ threshold | 49.2% | 60.6% (53.7%,64.5%) | 52.8% (43.9%,59.9%) | 178.8 (107.1,268.1) | 168.4 (98.6,242.1) |
|  | Full sharing | 49.1% | 59.4% (52.2%,63.9%) | 21.6% (20.4%,22.7%) | 171.0 (109.2,259.6) | 70.0 (64.3,77.4) |
| Low income | Default | 1.7% | 72.6% (63.9%,77.0%) | 72.6% (63.9%,77.0%) | 126.4 (72.3,201.9) | 126.4 (72.3,201.9) |
|  | 2 dose threshold | 11.6% | 72.5% (63.7%,77.0%) | 72.5% (63.7%,77.0%) | 126.1 (72.3,201.5) | 126.1 (72.3,201.5) |
|  | 40+ threshold | 26.7% | 71.9% (62.2%,76.8%) | 71.9% (62.2%,76.8%) | 119.2 (69.8,190.4) | 119.3 (69.8,190.8) |
|  | 65+ threshold | 38.5% | 68.3% (55.1%,74.9%) | 56.8% (44.2%,66.4%) | 94.7 (57.1,149.9) | 89.5 (53.3,145.8) |
|  | Full sharing | 40.6% | 63.5% (49.5%,72.0%) | 22.5% (20.7%,24.1%) | 71.8 (48.7,107.1) | 30.7 (27.4,35.2) |
| Lower middle income | Default | 18.4% | 82.2% (75.3%,86.3%) | 82.2% (75.3%,86.3%) | 193.4 (164.0,230.9) | 193.4 (164.0,230.9) |
|  | 2 dose threshold | 28.4% | 82.1% (75.2%,86.3%) | 82.1% (75.2%,86.3%) | 193.6 (164.1,230.9) | 193.6 (164.1,230.9) |
|  | 40+ threshold | 39.2% | 81.8% (74.3%,86.2%) | 81.8% (74.3%,86.2%) | 190.2 (161.2,227.1) | 190.3 (161.2,227.2) |
|  | 65+ threshold | 46.0% | 80.4% (71.4%,85.4%) | 76.1% (65.7%,82.8%) | 171.7 (144.6,205.0) | 167.6 (141.8,200.8) |
|  | Full sharing | 47.3% | 78.6% (69.0%,84.3%) | 29.9% (27.3%,32.4%) | 146.3 (123.7,172.6) | 65.1 (56.2,73.7) |
| Higher middle income | Default | 65.9% | 40.7% (38.3%,42.1%) | 40.7% (38.3%,42.1%) | 168.9 (63.1,269.7) | 168.9 (63.1,269.7) |
|  | 2 dose threshold | 65.9% | 40.7% (38.3%,42.1%) | 40.7% (38.3%,42.1%) | 174.7 (80.6,269.7) | 174.7 (80.6,269.7) |
|  | 40+ threshold | 56.3% | 40.6% (38.1%,42.1%) | 40.6% (38.1%,42.1%) | 172.5 (79.7,265.9) | 172.5 (79.8,266.2) |
|  | 65+ threshold | 51.3% | 40.0% (36.8%,41.8%) | 32.8% (27.0%,37.7%) | 161.0 (74.3,247.7) | 149.8 (69.4,227.8) |
|  | Full sharing | 51.2% | 39.4% (35.5%,41.6%) | 15.5% (14.4%,16.6%) | 150.5 (63.6,236.2) | 63.8 (51.9,80.8) |
| High income | Default | 72.2% | 50.8% (40.1%,59.0%) | 50.8% (40.1%,59.0%) | 237.7 (124.8,444.9) | 237.7 (124.8,444.9) |
|  | 2 dose threshold | 77.8% | 50.7% (40.0%,58.8%) | 50.5% (39.6%,58.8%) | 253.8 (161.1,444.8) | 253.8 (161.0,444.7) |
|  | 40+ threshold | 68.2% | 56.4% (47.1%,62.9%) | 47.9% (36.4%,57.8%) | 257.3 (166.2,440.0) | 249.7 (157.1,436.7) |
|  | 65+ threshold | 57.0% | 60.1% (52.1%,66.2%) | 43.9% (32.3%,56.3%) | 253.3 (167.7,370.9) | 230.2 (143.7,341.2) |
|  | Full sharing | 52.8% | 60.5% (53.2%,66.7%) | 16.3% (15.5%,17.1%) | 296.2 (203.2,418.4) | 105.1 (97.0,113.6) |

What is notable about the later increases in infection is that the corresponding mortality is significantly less pronounced (Fig. 3, middle row). This is due to the majority of vulnerable people having been offered vaccination in high income nations by this time in all scenarios, and hence the bulk of this increased infection would be felt by the younger and less vulnerable. Conversely, the earlier infection prevention in lower income nations by increased vaccine sharing is likely to have had included many of the elderly and vulnerable, and as such translated into a significant saving of lives. As a result of this we estimate there would have been 22 fewer deaths per 100,000 globally in the full sharing scenario, with lower income countries again seeing the greatest benefits with 40 fewer deaths per 100,000 (Fig. 3, bottom row). The bulk of these benefits are not seen in later sharing scenarios however, with relatively few countries reaching the 40+ age threshold before mid to late 2021 – by which time many low income countries have also already experienced large amounts of infection. Hence, the large gains are only seen in earlier sharing scenarios (65+ age threshold/full sharing), though in the late stages of 2021 some evidence is seen of potential further benefits in all scenarios emerging as new variants drive transmission increases and vaccines begin to wane.

If the increased infection seen in high income countries due to increased vaccine sharing resulted in extended behavioural caution (adapted behaviour), we predict a much greater reduction in global infections (pale bars Fig. 3, bottom row). We find estimates of global population infected and mortality rates by the end of 2021 were dramatically cut for the full vaccine sharing strategy scenario (21% infected, 47 deaths per 100,000) compared to the default scenario (58% infected, 122 deaths per 100,000). This is driven by the fact that the benefits of early vaccine sharing for lower income countries persist throughout 2021, reducing infection and hence the potential for the emergence and spread of variants. This creates a positive feedback loop with less infection causing less rise in transmission, itself leading to less infection; for infection levels below 30% we do not have the large jump in transmission that is associated with the Delta variant (Fig. 1).

## Discussion

We show that a more equitable approach to global distribution of vaccine over the course of 2021 would have reduced the level of global mortality associated with COVID-19 disease. Our conclusions are based on simulations fitted to historical infection and mortality estimates, and from varying the distribution of vaccination between countries while maintaining total vaccine supply. We build upon the findings of Wagner et al [21], extending their two-country conceptual model to a fitted simulation across 152 nations. In reality, any such historical deviation would likely precipitate a range of consequences, including changes in other policy areas, social behaviour, overall vaccine uptake, patterns of viral spread and variant evolution. While such confounding factors are difficult to predict, the simulation results presented here include potential changes to policy (or behaviour) and account for the consequences for variant accretion, increasing the robustness of our findings.

We predict the greatest advantages are associated with vaccine sharing earlier in the pandemic, with less extensive or delayed strategies presenting more modest savings. Due to the relative high transmissibility of SARS-COV-2 infection, countries without high and early vaccine coverage are likely to rapidly incur high infection levels and hence substantial immunity, as such the effects of late vaccine sharing to these countries are reduced. In addition, due to limited delivery capacities and increasing public distrust of vaccination [39], starting vaccination earlier may lead to more successful vaccination campaigns in general. However, with increasing transmission and possible immune escape from new variants [22] and the risk of waning efficacy, vaccine sharing remains highly important — our model suggests even late vaccine sharing once countries have delivered all second doses would see sizeable benefits as we move into 2022.

Our results suggest that increased vaccine sharing would have provided large benefits in low and lower-middle income countries; however, this comes at a cost to some high income countries where increases or prolonging of NPI measures would be required to suppress disease in the short term. Since increased sharing distributes disease burden more equitably, it could also have helped prevent any one country from experiencing an overwhelming and unmanageable wave of disease, and reduced the impact in the poorest nations that are least well equipped to manage the pandemic. In addition, as higher sharing scenarios delay infections until later in the year, these infections would have occurred once knowledge and treatments had improved and so been better managed. Furthermore, in this analysis we have assumed that vaccine efficacy profiles used are uniform across nations, however many lower income countries rely on substantially less effective vaccines than the more desirable counterparts employed by high income nations [32]. This imbalance means that the heterogeneity in effective vaccine coverage may effectively be even greater than assumed and increasing vaccine sharing to address the imbalance even more critical.

We have concentrated on supply constraints; assuming that a fixed amount of vaccine has been available throughout 2021, the key issue addressed is where this should be deployed. Supply has historically been the most significant factor causing heterogeneity in worldwide coverage [40] — when vaccines first became available the limited quantities produced were primarily purchased by wealthier nations. A confounding factor in this calculation is that since the nations producing and financing the vaccines have typically had access to a large amounts of vaccine, there has been little incentive to increase production [41]. One might hypothesise that increased sharing might have encouraged additional resources being put into production, increasing the overall volume of vaccines available.

More recently, with a number of different vaccines now being produced and the success of the COVAX scheme increasing vaccine availability [7], limitations surrounding delivery and uptake are becoming increasingly important [42]. Many lower income countries lack the infrastructure needed to rapidly deliver vaccines on the scale required, especially where there are large, hard to reach population sectors. Similarly, while vaccine hesitancy has been a recognised problem in all nations, in countries where public health messaging and education is limited, hesitancy is becoming a severe limiting factor for increased vaccine coverage [43, 39, 44]. Future support may therefore need to be more than prevision of vaccine doses, and may need to include assistance with vaccine delivery and logistical support.

Vaccines generally offer greater protection against severe disease than infection [14, 45], and the effects against severe disease are likely to be more robust against both waning immunity and vaccine escape [14, 46]. Hence, deploying vaccines to regions where there remains a high proportion of unprotected vulnerable individuals has a much greater impact per-dose than extending vaccination in nations that have already protected the majority of their vulnerable population. A complication to this vaccine equity picture is that the number of elderly and vulnerable individuals is larger in high-income countries, so even when vaccine is distributed purely according to need, heterogeneities will still exist.

Given its lack of significant impact during 2021, we have not explored the major threat of variants that escape vaccine and/or naturally induced immunity [47]. The emergence of Omicron in November 2021 poses just such a threat, with the potential for large waves of infection and the need to re-vaccinate some vulnerable populations. This recent emergence strengthens the arguments for vaccine sharing as a means of reducing the global levels of infection and hence retarding the accumulation of new variants.

Our model-based results reinforce the global public health message that vaccine nationalism (protecting one’s own country to the detriment of others) not only leads to greater levels of infection and mortality worldwide, but also adversely impacts all nations in the longer term [48, 49, 50].

## Supporting information

Full country table

## Data Availability

The study was based on data from a variety of publicly available sources: population demographic data provided by the WHO [26]; Income group classifications given by the World Bank [24]; COVID vaccine deployment provided by Our World in Data [6]; COVID mortality and infection estimates to date made by the Institute for Health Metrics and Evaluation [19]; data on COVID variants collated by GISAID
[21].

## Author contributions

**Conceptualisation:** Sam Moore; Matt J. Keeling.

**Formal analysis:** Sam Moore.

**Investigation:** Sam Moore.

**Methodology:** Sam Moore; Matt J. Keeling.

**Software:** Sam Moore.

**Validation:** Sam Moore; Matt J. Keeling; Edward M. Hill; Louise Dyson; Michael J. Tildesley.

**Visualisation:** Sam Moore.

**Writing - original draft:** Sam Moore.

**Writing - review & editing:** Sam Moore; Matt J. Keeling; Edward M. Hill; Louise Dyson; Michael J. Tildesley.

## Data sharing statement

The study was based on data from a variety of publicly available sources: population demographic data provided by the WHO [26]; Income group classifications given by the World Bank [24]; COVID vaccine deployment provided by Our World in Data [5]; COVID mortality and infection estimates to date made by the Institute for Health Metrics and Evaluation [18]; data on COVID variants collated by GISAID [20].

