## Supplementary material for "The impacts of increased global vaccine sharing on the COVID-19 pandemic; a retrospective modelling study": Full country table

### Supplementary Information, full results table: The impacts of increased global vaccine sharing on the COVID-19 pandemic; a retrospective modelling study

**Sam Moore, Edward M. Hill, Louise Dyson, Michael J. Tildesley, Matt J. Keeling**

| Country | Income bracket | Strategy | Proportion vaccinated | Proportion infected | | Mortalities per 100,000 | |
|---|---|---|---|---|---|---|---|
| | | | | unchanged | adapted | unchanged | adapted |
| Afghanistan | Low income | Default | 0.03 | 0.95 (0.90,0.97) | 0.95 (0.90,0.97) | 312 (167,489) | 312 (167,489) |
| Afghanistan | Low income | 2 dose threshold | 0.12 | 0.95 (0.90,0.97) | 0.95 (0.90,0.97) | 312 (167,488) | 312 (167,488) |
| Afghanistan | Low income | 40+ threshold | 0.27 | 0.95 (0.90,0.96) | 0.95 (0.90,0.96) | 303 (162,471) | 303 (162,471) |
| Afghanistan | Low income | 65+ threshold | 0.39 | 0.93 (0.89,0.96) | 0.83 (0.61,0.93) | 236 (139,369) | 216 (134,341) |
| Afghanistan | Low income | Full sharing | 0.41 | 0.92 (0.86,0.95) | 0.36 (0.30,0.42) | 185 (112,281) | 103 (70,142) |
| Angola | Lower middle income | Default | 0.04 | 0.76 (0.64,0.84) | 0.76 (0.64,0.84) | 109 (64,168) | 109 (64,168) |
| Angola | Lower middle income | 2 dose threshold | 0.13 | 0.76 (0.64,0.84) | 0.76 (0.64,0.84) | 109 (64,167) | 109 (64,167) |
| Angola | Lower middle income | 40+ threshold | 0.26 | 0.76 (0.62,0.84) | 0.76 (0.62,0.84) | 105 (61,163) | 106 (61,163) |
| Angola | Lower middle income | 65+ threshold | 0.36 | 0.73 (0.56,0.83) | 0.59 (0.38,0.78) | 82 (48,128) | 72 (40,122) |
| Angola | Lower middle income | Full sharing | 0.38 | 0.67 (0.49,0.79) | 0.20 (0.14,0.25) | 60 (38,90) | 23 (17,28) |
| Albania | Upper middle income | Default | 0.28 | 0.84 (0.79,0.87) | 0.84 (0.79,0.87) | 343 (225,510) | 343 (225,510) |
| Albania | Upper middle income | 2 dose threshold | 0.38 | 0.84 (0.79,0.87) | 0.84 (0.79,0.87) | 343 (225,510) | 343 (225,510) |
| Albania | Upper middle income | 40+ threshold | 0.52 | 0.83 (0.79,0.86) | 0.83 (0.79,0.86) | 342 (225,509) | 342 (225,509) |
| Albania | Upper middle income | 65+ threshold | 0.55 | 0.83 (0.79,0.86) | 0.81 (0.77,0.85) | 341 (224,506) | 340 (224,506) |
| Albania | Upper middle income | Full sharing | 0.52 | 0.83 (0.79,0.86) | 0.49 (0.40,0.55) | 321 (212,476) | 198 (145,259) |
| United Arab Emirates | High income | Default | 1.06 | 0.45 (0.34,0.55) | 0.45 (0.34,0.55) | 60 (38,85) | 60 (38,85) |
| United Arab Emirates | High income | 2 dose threshold | 0.99 | 0.45 (0.34,0.55) | 0.45 (0.34,0.55) | 60 (38,85) | 60 (38,85) |
| United Arab Emirates | High income | 40+ threshold | 0.50 | 0.75 (0.72,0.78) | 0.53 (0.37,0.61) | 99 (54,158) | 71 (40,111) |
| United Arab Emirates | High income | 65+ threshold | 0.55 | 0.77 (0.74,0.79) | 0.53 (0.43,0.61) | 128 (65,211) | 92 (50,143) |
| United Arab Emirates | High income | Full sharing | 0.55 | 0.75 (0.71,0.76) | 0.15 (0.12,0.17) | 165 (84,272) | 36 (24,52) |
| Argentina | Upper middle income | Default | 0.57 | 0.86 (0.80,0.89) | 0.86 (0.80,0.89) | 303 (247,367) | 303 (247,367) |
| Argentina | Upper middle income | 2 dose threshold | 0.63 | 0.86 (0.80,0.89) | 0.86 (0.80,0.89) | 303 (247,367) | 303 (247,367) |
| Argentina | Upper middle income | 40+ threshold | 0.53 | 0.86 (0.80,0.89) | 0.86 (0.80,0.89) | 302 (245,365) | 302 (245,366) |
| Argentina | Upper middle income | 65+ threshold | 0.51 | 0.85 (0.79,0.89) | 0.80 (0.66,0.89) | 293 (239,352) | 285 (230,351) |
| Argentina | Upper middle income | Full sharing | 0.49 | 0.86 (0.79,0.89) | 0.31 (0.27,0.35) | 299 (244,362) | 130 (114,148) |
| Armenia | Upper middle income | Default | 0.03 | 0.90 (0.85,0.94) | 0.90 (0.85,0.94) | 390 (262,558) | 390 (262,558) |

| Country | Income bracket | Strategy | Proportion vaccinated | Proportion infected | | Mortalities per 100,000 | |
|---|---|---|---|---|---|---|---|
| | | | | unchanged | adapted | unchanged | adapted |
| Armenia | Upper middle income | 2 dose threshold | 0.14 | 0.90 (0.85,0.94) | 0.90 (0.85,0.94) | 390 (262,557) | 390 (262,557) |
| Armenia | Upper middle income | 40+ threshold | 0.33 | 0.90 (0.85,0.94) | 0.90 (0.85,0.94) | 386 (260,550) | 386 (260,551) |
| Armenia | Upper middle income | 65+ threshold | 0.48 | 0.89 (0.84,0.94) | 0.89 (0.83,0.94) | 364 (239,519) | 363 (238,518) |
| Armenia | Upper middle income | Full sharing | 0.51 | 0.88 (0.83,0.93) | 0.62 (0.59,0.66) | 313 (210,451) | 244 (174,339) |
| Australia | High income | Default | 0.35 | 0.07 (0.01,0.18) | 0.07 (0.01,0.18) | 27 (12,54) | 27 (12,54) |
| Australia | High income | 2 dose threshold | 0.45 | 0.06 (0.01,0.18) | 0.06 (0.01,0.18) | 26 (12,54) | 26 (12,54) |
| Australia | High income | 40+ threshold | 0.56 | 0.06 (0.01,0.17) | 0.06 (0.01,0.17) | 24 (12,47) | 24 (12,47) |
| Australia | High income | 65+ threshold | 0.53 | 0.05 (0.01,0.13) | 0.01 (0.01,0.02) | 18 (10,31) | 12 (9,18) |
| Australia | High income | Full sharing | 0.51 | 0.05 (0.01,0.14) | 0.01 (0.01,0.01) | 18 (9,33) | 7 (6,8) |
| Austria | High income | Default | 0.80 | 0.46 (0.33,0.54) | 0.46 (0.33,0.54) | 251 (181,320) | 251 (181,320) |
| Austria | High income | 2 dose threshold | 0.84 | 0.46 (0.33,0.54) | 0.46 (0.33,0.54) | 251 (181,320) | 251 (181,320) |
| Austria | High income | 40+ threshold | 0.73 | 0.46 (0.33,0.54) | 0.46 (0.32,0.54) | 250 (181,320) | 250 (181,320) |
| Austria | High income | 65+ threshold | 0.60 | 0.48 (0.35,0.54) | 0.40 (0.28,0.51) | 252 (186,316) | 241 (176,310) |
| Austria | High income | Full sharing | 0.53 | 0.50 (0.37,0.56) | 0.16 (0.14,0.19) | 305 (216,404) | 118 (96,134) |
| Azerbaijan | Upper middle income | Default | 0.34 | 0.85 (0.80,0.89) | 0.85 (0.80,0.89) | 269 (174,387) | 269 (174,387) |
| Azerbaijan | Upper middle income | 2 dose threshold | 0.42 | 0.85 (0.80,0.89) | 0.85 (0.80,0.89) | 269 (174,387) | 269 (174,387) |
| Azerbaijan | Upper middle income | 40+ threshold | 0.50 | 0.84 (0.80,0.89) | 0.84 (0.80,0.89) | 268 (174,386) | 268 (174,386) |
| Azerbaijan | Upper middle income | 65+ threshold | 0.49 | 0.84 (0.80,0.89) | 0.81 (0.72,0.89) | 265 (171,380) | 261 (171,368) |
| Azerbaijan | Upper middle income | Full sharing | 0.50 | 0.84 (0.80,0.89) | 0.50 (0.45,0.58) | 256 (170,366) | 161 (118,207) |
| Belgium | High income | Default | 0.95 | 0.58 (0.45,0.64) | 0.58 (0.45,0.64) | 526 (408,677) | 526 (408,677) |
| Belgium | High income | 2 dose threshold | 0.94 | 0.58 (0.45,0.64) | 0.58 (0.45,0.64) | 526 (408,677) | 526 (408,677) |
| Belgium | High income | 40+ threshold | 0.71 | 0.58 (0.45,0.64) | 0.58 (0.45,0.64) | 525 (407,675) | 525 (407,676) |
| Belgium | High income | 65+ threshold | 0.58 | 0.59 (0.46,0.64) | 0.51 (0.37,0.63) | 525 (408,669) | 510 (396,658) |
| Belgium | High income | Full sharing | 0.52 | 0.60 (0.47,0.66) | 0.22 (0.20,0.26) | 622 (483,815) | 268 (227,317) |
| Benin | Lower middle income | Default | 0.00 | 0.52 (0.31,0.66) | 0.52 (0.31,0.66) | 86 (50,129) | 86 (50,129) |
| Benin | Lower middle income | 2 dose threshold | 0.10 | 0.52 (0.31,0.66) | 0.52 (0.31,0.66) | 86 (50,129) | 86 (50,129) |
| Benin | Lower middle income | 40+ threshold | 0.25 | 0.51 (0.30,0.65) | 0.51 (0.30,0.65) | 73 (43,112) | 73 (43,112) |
| Benin | Lower middle income | 65+ threshold | 0.38 | 0.38 (0.20,0.49) | 0.25 (0.14,0.35) | 42 (27,63) | 38 (25,58) |
| Benin | Lower middle income | Full sharing | 0.41 | 0.30 (0.16,0.40) | 0.07 (0.05,0.10) | 33 (22,48) | 10 (7,13) |
| Burkina Faso | Low income | Default | 0.00 | 0.69 (0.55,0.78) | 0.69 (0.55,0.78) | 47 (27,75) | 47 (27,75) |
| Burkina Faso | Low income | 2 dose threshold | 0.10 | 0.68 (0.55,0.77) | 0.68 (0.55,0.77) | 46 (26,75) | 46 (26,75) |
| Burkina Faso | Low income | 40+ threshold | 0.25 | 0.65 (0.53,0.74) | 0.65 (0.53,0.74) | 43 (25,72) | 43 (25,72) |
| Burkina Faso | Low income | 65+ threshold | 0.37 | 0.61 (0.49,0.72) | 0.60 (0.48,0.72) | 40 (23,67) | 40 (23,67) |
| Burkina Faso | Low income | Full sharing | 0.39 | 0.60 (0.48,0.71) | 0.34 (0.22,0.44) | 36 (21,60) | 18 (13,25) |
| Bangladesh | Lower middle income | Default | 0.05 | 0.81 (0.76,0.85) | 0.81 (0.76,0.85) | 93 (53,142) | 93 (53,142) |

| Country | Income bracket | Strategy | Proportion vaccinated | Proportion infected | | Mortalities per 100,000 | |
|---|---|---|---|---|---|---|---|
| | | | | unchanged | adapted | unchanged | adapted |
| Bangladesh | Lower middle income | 2 dose threshold | 0.16 | 0.81 (0.76,0.85) | 0.81 (0.76,0.85) | 93 (52,142) | 93 (52,142) |
| Bangladesh | Lower middle income | 40+ threshold | 0.34 | 0.80 (0.76,0.85) | 0.80 (0.76,0.84) | 92 (52,139) | 92 (52,139) |
| Bangladesh | Lower middle income | 65+ threshold | 0.48 | 0.79 (0.75,0.83) | 0.72 (0.57,0.82) | 86 (48,129) | 82 (45,127) |
| Bangladesh | Lower middle income | Full sharing | 0.49 | 0.79 (0.75,0.83) | 0.36 (0.26,0.45) | 76 (44,113) | 43 (30,55) |
| Bulgaria | Upper middle income | Default | 0.19 | 0.83 (0.78,0.85) | 0.83 (0.78,0.85) | 371 (246,556) | 371 (246,556) |
| Bulgaria | Upper middle income | 2 dose threshold | 0.30 | 0.83 (0.78,0.85) | 0.83 (0.78,0.85) | 371 (246,556) | 371 (246,556) |
| Bulgaria | Upper middle income | 40+ threshold | 0.47 | 0.82 (0.77,0.85) | 0.82 (0.77,0.85) | 370 (245,554) | 370 (245,555) |
| Bulgaria | Upper middle income | 65+ threshold | 0.56 | 0.82 (0.77,0.85) | 0.80 (0.73,0.85) | 365 (242,546) | 364 (240,545) |
| Bulgaria | Upper middle income | Full sharing | 0.53 | 0.82 (0.77,0.85) | 0.41 (0.32,0.49) | 358 (238,536) | 227 (172,297) |
| Bahrain | High income | Default | 0.94 | 0.80 (0.69,0.86) | 0.80 (0.69,0.86) | 125 (83,167) | 125 (83,167) |
| Bahrain | High income | 2 dose threshold | 0.93 | 0.80 (0.69,0.86) | 0.80 (0.69,0.86) | 125 (83,167) | 125 (83,167) |
| Bahrain | High income | 40+ threshold | 0.50 | 0.82 (0.74,0.87) | 0.56 (0.49,0.66) | 129 (87,174) | 97 (66,129) |
| Bahrain | High income | 65+ threshold | 0.52 | 0.90 (0.85,0.92) | 0.67 (0.57,0.73) | 221 (142,305) | 147 (93,208) |
| Bahrain | High income | Full sharing | 0.54 | 0.88 (0.82,0.91) | 0.26 (0.23,0.29) | 189 (125,260) | 63 (44,82) |
| Bosnia and Herzegovina | Upper middle income | Default | 0.13 | 0.84 (0.81,0.87) | 0.84 (0.81,0.87) | 306 (240,422) | 306 (240,422) |
| Bosnia and Herzegovina | Upper middle income | 2 dose threshold | 0.24 | 0.84 (0.81,0.87) | 0.84 (0.81,0.87) | 306 (240,422) | 306 (240,422) |
| Bosnia and Herzegovina | Upper middle income | 40+ threshold | 0.43 | 0.84 (0.81,0.87) | 0.84 (0.81,0.87) | 304 (238,419) | 304 (238,420) |
| Bosnia and Herzegovina | Upper middle income | 65+ threshold | 0.52 | 0.84 (0.80,0.87) | 0.82 (0.75,0.87) | 295 (227,405) | 294 (225,404) |
| Bosnia and Herzegovina | Upper middle income | Full sharing | 0.54 | 0.83 (0.79,0.87) | 0.50 (0.43,0.55) | 271 (210,365) | 189 (162,229) |
| Belarus | Upper middle income | Default | 0.15 | 0.79 (0.73,0.84) | 0.79 (0.73,0.84) | 534 (315,885) | 534 (315,885) |
| Belarus | Upper middle income | 2 dose threshold | 0.25 | 0.79 (0.73,0.84) | 0.79 (0.73,0.84) | 534 (315,885) | 534 (315,885) |
| Belarus | Upper middle income | 40+ threshold | 0.44 | 0.79 (0.73,0.84) | 0.79 (0.73,0.84) | 525 (312,868) | 525 (312,869) |
| Belarus | Upper middle income | 65+ threshold | 0.51 | 0.77 (0.70,0.83) | 0.76 (0.67,0.83) | 460 (282,727) | 456 (282,724) |
| Belarus | Upper middle income | Full sharing | 0.52 | 0.76 (0.68,0.82) | 0.27 (0.18,0.36) | 411 (254,637) | 168 (124,212) |
| Bolivia | Lower middle income | Default | 0.25 | 0.88 (0.85,0.90) | 0.88 (0.85,0.90) | 1025 (628,1471) | 1025 (628,1471) |
| Bolivia | Lower middle income | 2 dose threshold | 0.34 | 0.88 (0.85,0.90) | 0.88 (0.85,0.90) | 1025 (628,1470) | 1025 (628,1470) |
| Bolivia | Lower middle income | 40+ threshold | 0.42 | 0.88 (0.85,0.89) | 0.88 (0.85,0.90) | 1015 (620,1455) | 1015 (620,1456) |
| Bolivia | Lower middle income | 65+ threshold | 0.45 | 0.87 (0.84,0.89) | 0.85 (0.79,0.88) | 952 (564,1390) | 944 (559,1378) |
| Bolivia | Lower middle income | Full sharing | 0.47 | 0.86 (0.83,0.88) | 0.58 (0.53,0.62) | 805 (487,1169) | 504 (307,679) |
| Brazil | Upper middle income | Default | 0.54 | 0.82 (0.78,0.84) | 0.82 (0.78,0.84) | 245 (206,291) | 245 (206,291) |
| Brazil | Upper middle income | 2 dose threshold | 0.61 | 0.82 (0.78,0.84) | 0.82 (0.78,0.84) | 245 (206,291) | 245 (206,291) |
| Brazil | Upper middle income | 40+ threshold | 0.55 | 0.82 (0.78,0.84) | 0.82 (0.78,0.84) | 245 (205,290) | 245 (206,290) |
| Brazil | Upper middle income | 65+ threshold | 0.52 | 0.82 (0.78,0.84) | 0.72 (0.63,0.80) | 242 (204,287) | 231 (190,279) |
| Brazil | Upper middle income | Full sharing | 0.51 | 0.82 (0.77,0.84) | 0.35 (0.32,0.39) | 239 (202,283) | 115 (105,127) |
| Barbados | High income | Default | 0.48 | 0.51 (0.36,0.62) | 0.51 (0.36,0.62) | 76 (62,96) | 76 (62,96) |

| Country | Income bracket | Strategy | Proportion vaccinated | Proportion infected | | Mortalities per 100,000 | |
|---|---|---|---|---|---|---|---|
| | | | | unchanged | adapted | unchanged | adapted |
| Barbados | High income | 2 dose threshold | 0.56 | 0.50 (0.36,0.62) | 0.50 (0.36,0.62) | 76 (62,96) | 76 (62,96) |
| Barbados | High income | 40+ threshold | 0.64 | 0.47 (0.32,0.60) | 0.47 (0.31,0.60) | 74 (61,93) | 74 (61,93) |
| Barbados | High income | 65+ threshold | 0.58 | 0.53 (0.39,0.65) | 0.19 (0.14,0.24) | 83 (68,104) | 59 (43,85) |
| Barbados | High income | Full sharing | 0.53 | 0.59 (0.45,0.70) | 0.13 (0.10,0.22) | 102 (82,130) | 40 (23,88) |
| Central African Republic | Low income | Default | 0.02 | 0.84 (0.72,0.90) | 0.84 (0.72,0.90) | 226 (124,338) | 226 (124,338) |
| Central African Republic | Low income | 2 dose threshold | 0.11 | 0.84 (0.72,0.90) | 0.84 (0.72,0.90) | 226 (124,338) | 226 (124,338) |
| Central African Republic | Low income | 40+ threshold | 0.26 | 0.83 (0.72,0.90) | 0.84 (0.72,0.90) | 221 (121,330) | 222 (121,330) |
| Central African Republic | Low income | 65+ threshold | 0.38 | 0.83 (0.70,0.90) | 0.83 (0.70,0.90) | 194 (105,293) | 194 (105,291) |
| Central African Republic | Low income | Full sharing | 0.40 | 0.80 (0.66,0.89) | 0.39 (0.29,0.49) | 141 (81,207) | 72 (52,95) |
| Canada | High income | Default | 1.02 | 0.53 (0.29,0.65) | 0.53 (0.29,0.65) | 188 (130,275) | 188 (130,275) |
| Canada | High income | 2 dose threshold | 0.97 | 0.53 (0.29,0.65) | 0.53 (0.29,0.65) | 188 (130,274) | 188 (130,274) |
| Canada | High income | 40+ threshold | 0.71 | 0.53 (0.29,0.65) | 0.53 (0.29,0.65) | 187 (129,273) | 187 (129,273) |
| Canada | High income | 65+ threshold | 0.57 | 0.57 (0.34,0.66) | 0.39 (0.21,0.55) | 194 (136,279) | 173 (122,245) |
| Canada | High income | Full sharing | 0.53 | 0.59 (0.35,0.68) | 0.14 (0.09,0.18) | 231 (157,356) | 74 (59,88) |
| Switzerland | High income | Default | 0.75 | 0.48 (0.39,0.55) | 0.48 (0.39,0.55) | 579 (232,1517) | 579 (232,1517) |
| Switzerland | High income | 2 dose threshold | 0.80 | 0.48 (0.39,0.55) | 0.48 (0.39,0.55) | 579 (232,1517) | 579 (232,1517) |
| Switzerland | High income | 40+ threshold | 0.71 | 0.48 (0.39,0.55) | 0.48 (0.39,0.55) | 576 (231,1509) | 577 (231,1512) |
| Switzerland | High income | 65+ threshold | 0.58 | 0.49 (0.40,0.55) | 0.38 (0.30,0.47) | 571 (231,1468) | 528 (217,1318) |
| Switzerland | High income | Full sharing | 0.53 | 0.51 (0.42,0.58) | 0.17 (0.14,0.21) | 703 (269,1924) | 197 (126,304) |
| Chile | High income | Default | 1.02 | 0.64 (0.50,0.71) | 0.64 (0.50,0.71) | 263 (224,300) | 263 (224,300) |
| Chile | High income | 2 dose threshold | 0.91 | 0.64 (0.50,0.71) | 0.64 (0.50,0.71) | 263 (224,300) | 263 (224,300) |
| Chile | High income | 40+ threshold | 0.63 | 0.69 (0.59,0.74) | 0.50 (0.35,0.66) | 268 (232,301) | 251 (212,295) |
| Chile | High income | 65+ threshold | 0.55 | 0.83 (0.78,0.85) | 0.41 (0.34,0.49) | 314 (274,345) | 243 (211,283) |
| Chile | High income | Full sharing | 0.52 | 0.84 (0.79,0.87) | 0.18 (0.16,0.20) | 352 (305,394) | 104 (100,109) |
| China | Upper middle income | Default | 1.03 | 0.00 (0.00,0.00) | 0.00 (0.00,0.00) | 1 (1,1) | 1 (1,1) |
| China | Upper middle income | 2 dose threshold | 0.93 | 0.00 (0.00,0.00) | 0.00 (0.00,0.00) | 1 (1,1) | 1 (1,1) |
| China | Upper middle income | 40+ threshold | 0.67 | 0.00 (0.00,0.00) | 0.00 (0.00,0.00) | 1 (1,1) | 1 (1,1) |
| China | Upper middle income | 65+ threshold | 0.53 | 0.00 (0.00,0.00) | 0.00 (0.00,0.00) | 1 (1,1) | 1 (1,1) |
| China | Upper middle income | Full sharing | 0.53 | 0.00 (0.00,0.00) | 0.00 (0.00,0.00) | 1 (1,1) | 0 (0,0) |
| Cameroon | Lower middle income | Default | 0.01 | 0.71 (0.60,0.77) | 0.71 (0.60,0.77) | 82 (39,135) | 82 (39,135) |
| Cameroon | Lower middle income | 2 dose threshold | 0.11 | 0.71 (0.60,0.77) | 0.71 (0.60,0.77) | 82 (39,135) | 82 (39,135) |
| Cameroon | Lower middle income | 40+ threshold | 0.26 | 0.70 (0.59,0.77) | 0.70 (0.59,0.77) | 79 (38,132) | 79 (38,132) |
| Cameroon | Lower middle income | 65+ threshold | 0.39 | 0.68 (0.54,0.76) | 0.66 (0.50,0.77) | 67 (30,115) | 66 (30,115) |
| Cameroon | Lower middle income | Full sharing | 0.41 | 0.64 (0.49,0.73) | 0.25 (0.19,0.35) | 51 (25,83) | 23 (16,29) |
| Democratic Republic of the Congo | Low income | Default | 0.00 | 0.72 (0.55,0.80) | 0.72 (0.55,0.80) | 102 (65,158) | 102 (65,158) |

| Country | Income bracket | Strategy | Proportion vaccinated | Proportion infected | | Mortalities per 100,000 | |
|---|---|---|---|---|---|---|---|
| | | | | unchanged | adapted | unchanged | adapted |
| Democratic Republic of the Congo | Low income | 2 dose threshold | 0.09 | 0.72 (0.55,0.80) | 0.72 (0.55,0.80) | 101 (65,158) | 101 (65,158) |
| Democratic Republic of the Congo | Low income | 40+ threshold | 0.24 | 0.71 (0.55,0.80) | 0.71 (0.54,0.80) | 92 (59,142) | 92 (59,142) |
| Democratic Republic of the Congo | Low income | 65+ threshold | 0.36 | 0.64 (0.44,0.75) | 0.44 (0.31,0.55) | 61 (40,88) | 54 (36,75) |
| Democratic Republic of the Congo | Low income | Full sharing | 0.38 | 0.56 (0.36,0.69) | 0.21 (0.16,0.26) | 50 (34,69) | 22 (16,29) |
| Colombia | Upper middle income | Default | 0.34 | 0.83 (0.75,0.87) | 0.83 (0.75,0.87) | 359 (280,453) | 359 (280,453) |
| Colombia | Upper middle income | 2 dose threshold | 0.43 | 0.83 (0.75,0.87) | 0.83 (0.75,0.87) | 359 (280,453) | 359 (280,453) |
| Colombia | Upper middle income | 40+ threshold | 0.50 | 0.83 (0.75,0.87) | 0.83 (0.75,0.87) | 355 (277,448) | 355 (277,448) |
| Colombia | Upper middle income | 65+ threshold | 0.50 | 0.81 (0.73,0.86) | 0.75 (0.58,0.86) | 322 (253,411) | 310 (233,404) |
| Colombia | Upper middle income | Full sharing | 0.51 | 0.80 (0.70,0.85) | 0.29 (0.25,0.34) | 282 (223,353) | 117 (101,134) |
| Comoros | Lower middle income | Default | 0.11 | 0.80 (0.75,0.84) | 0.80 (0.75,0.84) | 33 (20,49) | 33 (20,49) |
| Comoros | Lower middle income | 2 dose threshold | 0.21 | 0.80 (0.75,0.84) | 0.80 (0.75,0.84) | 33 (20,49) | 33 (20,49) |
| Comoros | Lower middle income | 40+ threshold | 0.32 | 0.79 (0.74,0.84) | 0.79 (0.74,0.84) | 33 (20,49) | 33 (20,49) |
| Comoros | Lower middle income | 65+ threshold | 0.40 | 0.79 (0.74,0.83) | 0.79 (0.73,0.83) | 32 (20,48) | 32 (20,48) |
| Comoros | Lower middle income | Full sharing | 0.42 | 0.79 (0.73,0.83) | 0.31 (0.27,0.38) | 28 (18,43) | 9 (7,12) |
| Cabo Verde | Lower middle income | Default | 0.00 | 0.84 (0.77,0.88) | 0.84 (0.77,0.88) | 164 (99,258) | 164 (99,258) |
| Cabo Verde | Lower middle income | 2 dose threshold | 0.11 | 0.84 (0.77,0.88) | 0.84 (0.77,0.88) | 163 (98,258) | 163 (98,258) |
| Cabo Verde | Lower middle income | 40+ threshold | 0.30 | 0.83 (0.76,0.88) | 0.83 (0.77,0.88) | 155 (94,249) | 156 (94,249) |
| Cabo Verde | Lower middle income | 65+ threshold | 0.45 | 0.82 (0.74,0.87) | 0.81 (0.73,0.87) | 128 (77,209) | 127 (76,208) |
| Cabo Verde | Lower middle income | Full sharing | 0.48 | 0.79 (0.71,0.85) | 0.41 (0.33,0.50) | 94 (57,150) | 47 (33,64) |
| Costa Rica | Upper middle income | Default | 0.49 | 0.76 (0.67,0.81) | 0.76 (0.67,0.81) | 128 (102,164) | 128 (102,164) |
| Costa Rica | Upper middle income | 2 dose threshold | 0.56 | 0.76 (0.67,0.81) | 0.76 (0.67,0.81) | 128 (102,164) | 128 (102,164) |
| Costa Rica | Upper middle income | 40+ threshold | 0.55 | 0.76 (0.66,0.81) | 0.76 (0.66,0.81) | 127 (101,163) | 127 (101,163) |
| Costa Rica | Upper middle income | 65+ threshold | 0.52 | 0.75 (0.64,0.80) | 0.64 (0.47,0.78) | 118 (95,149) | 111 (86,144) |
| Costa Rica | Upper middle income | Full sharing | 0.51 | 0.74 (0.64,0.80) | 0.26 (0.20,0.33) | 121 (96,155) | 51 (49,55) |
| Cuba | Upper middle income | Default | 0.36 | 0.57 (0.44,0.78) | 0.57 (0.44,0.78) | 220 (138,319) | 220 (138,319) |
| Cuba | Upper middle income | 2 dose threshold | 0.45 | 0.57 (0.44,0.78) | 0.57 (0.44,0.78) | 220 (137,319) | 220 (137,319) |
| Cuba | Upper middle income | 40+ threshold | 0.60 | 0.56 (0.43,0.78) | 0.56 (0.43,0.78) | 210 (124,316) | 211 (124,316) |
| Cuba | Upper middle income | 65+ threshold | 0.50 | 0.53 (0.36,0.78) | 0.49 (0.22,0.78) | 154 (62,282) | 148 (51,281) |
| Cuba | Upper middle income | Full sharing | 0.53 | 0.48 (0.28,0.75) | 0.01 (0.01,0.02) | 102 (38,197) | 3 (2,5) |
| Cyprus | High income | Default | 0.57 | 0.51 (0.28,0.76) | 0.51 (0.28,0.76) | 86 (46,135) | 86 (46,135) |
| Cyprus | High income | 2 dose threshold | 0.64 | 0.51 (0.28,0.76) | 0.51 (0.28,0.76) | 86 (46,135) | 86 (46,135) |
| Cyprus | High income | 40+ threshold | 0.65 | 0.51 (0.27,0.76) | 0.51 (0.28,0.76) | 85 (45,135) | 85 (45,135) |
| Cyprus | High income | 65+ threshold | 0.56 | 0.51 (0.30,0.75) | 0.48 (0.19,0.75) | 81 (45,129) | 79 (40,128) |
| Cyprus | High income | Full sharing | 0.53 | 0.52 (0.30,0.77) | 0.11 (0.09,0.15) | 101 (54,161) | 22 (19,26) |
| Czech Republic | High income | Default | 0.71 | 0.76 (0.70,0.82) | 0.76 (0.70,0.82) | 233 (168,317) | 233 (168,317) |

| Country | Income bracket | Strategy | Proportion vaccinated | Proportion infected | | Mortalities per 100,000 | |
|---|---|---|---|---|---|---|---|
| | | | | unchanged | adapted | unchanged | adapted |
| Czech Republic | High income | 2 dose threshold | 0.76 | 0.76 (0.70,0.82) | 0.76 (0.70,0.82) | 233 (168,317) | 233 (168,317) |
| Czech Republic | High income | 40+ threshold | 0.71 | 0.76 (0.70,0.82) | 0.76 (0.70,0.82) | 232 (168,317) | 232 (168,317) |
| Czech Republic | High income | 65+ threshold | 0.58 | 0.77 (0.70,0.82) | 0.75 (0.67,0.83) | 232 (168,316) | 231 (168,316) |
| Czech Republic | High income | Full sharing | 0.53 | 0.77 (0.70,0.83) | 0.42 (0.28,0.53) | 247 (177,338) | 147 (124,168) |
| Germany | High income | Default | 0.83 | 0.41 (0.20,0.55) | 0.41 (0.20,0.55) | 200 (125,293) | 200 (125,293) |
| Germany | High income | 2 dose threshold | 0.87 | 0.41 (0.20,0.55) | 0.41 (0.20,0.55) | 200 (125,292) | 200 (125,292) |
| Germany | High income | 40+ threshold | 0.75 | 0.41 (0.20,0.55) | 0.41 (0.20,0.55) | 199 (125,290) | 199 (125,291) |
| Germany | High income | 65+ threshold | 0.60 | 0.41 (0.21,0.54) | 0.38 (0.18,0.55) | 192 (127,277) | 187 (120,282) |
| Germany | High income | Full sharing | 0.53 | 0.43 (0.23,0.56) | 0.09 (0.08,0.09) | 239 (145,353) | 67 (56,80) |
| Djibouti | Lower middle income | Default | 0.03 | 0.83 (0.71,0.92) | 0.83 (0.71,0.92) | 330 (165,613) | 330 (165,613) |
| Djibouti | Lower middle income | 2 dose threshold | 0.14 | 0.83 (0.71,0.92) | 0.83 (0.71,0.92) | 330 (165,613) | 330 (165,613) |
| Djibouti | Lower middle income | 40+ threshold | 0.32 | 0.82 (0.70,0.92) | 0.82 (0.70,0.92) | 320 (160,599) | 321 (161,600) |
| Djibouti | Lower middle income | 65+ threshold | 0.45 | 0.81 (0.68,0.92) | 0.80 (0.67,0.92) | 290 (144,540) | 288 (144,536) |
| Djibouti | Lower middle income | Full sharing | 0.47 | 0.79 (0.65,0.91) | 0.37 (0.30,0.43) | 212 (108,381) | 94 (58,144) |
| Denmark | High income | Default | 0.86 | 0.39 (0.20,0.51) | 0.39 (0.20,0.51) | 159 (102,232) | 159 (102,232) |
| Denmark | High income | 2 dose threshold | 0.90 | 0.39 (0.20,0.51) | 0.39 (0.20,0.51) | 159 (102,232) | 159 (102,232) |
| Denmark | High income | 40+ threshold | 0.70 | 0.39 (0.20,0.51) | 0.39 (0.20,0.51) | 158 (101,231) | 158 (101,231) |
| Denmark | High income | 65+ threshold | 0.59 | 0.39 (0.20,0.50) | 0.26 (0.15,0.39) | 155 (103,220) | 136 (91,192) |
| Denmark | High income | Full sharing | 0.53 | 0.42 (0.21,0.54) | 0.09 (0.07,0.11) | 211 (131,317) | 63 (49,75) |
| Dominican Republic | Upper middle income | Default | 0.70 | 0.59 (0.44,0.68) | 0.59 (0.44,0.68) | 201 (131,276) | 201 (131,276) |
| Dominican Republic | Upper middle income | 2 dose threshold | 0.74 | 0.59 (0.44,0.68) | 0.59 (0.44,0.68) | 201 (131,276) | 201 (131,276) |
| Dominican Republic | Upper middle income | 40+ threshold | 0.52 | 0.58 (0.44,0.67) | 0.58 (0.43,0.68) | 200 (131,274) | 201 (131,274) |
| Dominican Republic | Upper middle income | 65+ threshold | 0.48 | 0.60 (0.45,0.69) | 0.45 (0.33,0.56) | 200 (130,273) | 187 (123,252) |
| Dominican Republic | Upper middle income | Full sharing | 0.48 | 0.59 (0.43,0.68) | 0.23 (0.17,0.30) | 177 (115,242) | 75 (53,97) |
| Algeria | Lower middle income | Default | 0.08 | 0.57 (0.33,0.71) | 0.57 (0.33,0.71) | 152 (85,231) | 152 (85,231) |
| Algeria | Lower middle income | 2 dose threshold | 0.19 | 0.57 (0.33,0.71) | 0.57 (0.33,0.71) | 152 (85,231) | 152 (85,231) |
| Algeria | Lower middle income | 40+ threshold | 0.34 | 0.57 (0.32,0.70) | 0.57 (0.32,0.70) | 144 (80,221) | 144 (80,221) |
| Algeria | Lower middle income | 65+ threshold | 0.45 | 0.51 (0.27,0.66) | 0.29 (0.18,0.49) | 107 (63,158) | 87 (56,133) |
| Algeria | Lower middle income | Full sharing | 0.46 | 0.47 (0.23,0.63) | 0.12 (0.10,0.15) | 89 (55,129) | 36 (28,44) |
| Ecuador | Upper middle income | Default | 0.29 | 0.86 (0.84,0.88) | 0.86 (0.84,0.88) | 603 (391,866) | 603 (391,866) |
| Ecuador | Upper middle income | 2 dose threshold | 0.38 | 0.86 (0.84,0.88) | 0.86 (0.84,0.88) | 603 (391,866) | 603 (391,866) |
| Ecuador | Upper middle income | 40+ threshold | 0.45 | 0.86 (0.84,0.88) | 0.86 (0.84,0.88) | 599 (389,859) | 600 (389,860) |
| Ecuador | Upper middle income | 65+ threshold | 0.47 | 0.85 (0.83,0.87) | 0.83 (0.77,0.87) | 572 (370,835) | 568 (370,828) |
| Ecuador | Upper middle income | Full sharing | 0.48 | 0.84 (0.82,0.87) | 0.48 (0.43,0.51) | 501 (323,722) | 294 (208,394) |
| Egypt | Lower middle income | Default | 0.05 | 0.77 (0.70,0.86) | 0.77 (0.70,0.86) | 196 (89,324) | 196 (89,324) |

| Country | Income bracket | Strategy | Proportion vaccinated | Proportion infected | | Mortalities per 100,000 | |
|---|---|---|---|---|---|---|---|
| | | | | unchanged | adapted | unchanged | adapted |
| Egypt | Lower middle income | 2 dose threshold | 0.15 | 0.77 (0.70,0.86) | 0.77 (0.70,0.86) | 196 (89,324) | 196 (89,324) |
| Egypt | Lower middle income | 40+ threshold | 0.31 | 0.77 (0.70,0.86) | 0.77 (0.70,0.86) | 193 (88,318) | 193 (88,318) |
| Egypt | Lower middle income | 65+ threshold | 0.42 | 0.76 (0.66,0.85) | 0.74 (0.59,0.85) | 171 (82,287) | 169 (82,287) |
| Egypt | Lower middle income | Full sharing | 0.44 | 0.73 (0.62,0.84) | 0.39 (0.25,0.53) | 141 (70,225) | 76 (47,96) |
| Spain | High income | Default | 0.83 | 0.52 (0.42,0.60) | 0.52 (0.42,0.60) | 547 (409,672) | 547 (409,672) |
| Spain | High income | 2 dose threshold | 0.87 | 0.52 (0.42,0.60) | 0.52 (0.42,0.60) | 547 (409,672) | 547 (409,672) |
| Spain | High income | 40+ threshold | 0.73 | 0.52 (0.42,0.59) | 0.52 (0.42,0.59) | 546 (408,671) | 546 (408,671) |
| Spain | High income | 65+ threshold | 0.59 | 0.57 (0.47,0.65) | 0.39 (0.33,0.46) | 560 (419,686) | 523 (391,643) |
| Spain | High income | Full sharing | 0.53 | 0.60 (0.49,0.68) | 0.16 (0.15,0.18) | 663 (490,825) | 247 (206,283) |
| Estonia | High income | Default | 0.62 | 0.59 (0.42,0.73) | 0.59 (0.42,0.73) | 198 (129,292) | 198 (129,292) |
| Estonia | High income | 2 dose threshold | 0.68 | 0.59 (0.42,0.73) | 0.59 (0.42,0.73) | 198 (129,292) | 198 (129,292) |
| Estonia | High income | 40+ threshold | 0.70 | 0.59 (0.41,0.73) | 0.59 (0.41,0.73) | 197 (128,292) | 197 (128,292) |
| Estonia | High income | 65+ threshold | 0.59 | 0.60 (0.46,0.73) | 0.58 (0.36,0.73) | 194 (129,285) | 193 (124,285) |
| Estonia | High income | Full sharing | 0.52 | 0.62 (0.50,0.74) | 0.12 (0.10,0.14) | 241 (157,354) | 52 (40,67) |
| Ethiopia | Low income | Default | 0.02 | 0.71 (0.52,0.78) | 0.71 (0.52,0.78) | 144 (79,266) | 144 (79,266) |
| Ethiopia | Low income | 2 dose threshold | 0.13 | 0.71 (0.52,0.78) | 0.71 (0.52,0.78) | 144 (79,265) | 144 (79,265) |
| Ethiopia | Low income | 40+ threshold | 0.28 | 0.70 (0.51,0.78) | 0.70 (0.51,0.78) | 139 (77,255) | 139 (77,256) |
| Ethiopia | Low income | 65+ threshold | 0.40 | 0.68 (0.46,0.77) | 0.69 (0.45,0.78) | 115 (64,215) | 115 (64,214) |
| Ethiopia | Low income | Full sharing | 0.42 | 0.65 (0.41,0.76) | 0.20 (0.13,0.27) | 83 (49,147) | 29 (21,39) |
| Finland | High income | Default | 0.90 | 0.40 (0.24,0.51) | 0.40 (0.24,0.51) | 174 (92,329) | 174 (92,329) |
| Finland | High income | 2 dose threshold | 0.93 | 0.40 (0.24,0.51) | 0.40 (0.24,0.51) | 174 (92,329) | 174 (92,329) |
| Finland | High income | 40+ threshold | 0.73 | 0.40 (0.23,0.51) | 0.40 (0.24,0.51) | 174 (91,328) | 174 (91,328) |
| Finland | High income | 65+ threshold | 0.60 | 0.43 (0.29,0.52) | 0.35 (0.19,0.53) | 176 (96,320) | 167 (89,316) |
| Finland | High income | Full sharing | 0.52 | 0.47 (0.32,0.56) | 0.11 (0.07,0.15) | 225 (111,465) | 65 (44,92) |
| France | High income | Default | 0.79 | 0.64 (0.52,0.69) | 0.64 (0.52,0.69) | 288 (230,371) | 288 (230,371) |
| France | High income | 2 dose threshold | 0.83 | 0.64 (0.52,0.69) | 0.64 (0.52,0.69) | 288 (230,371) | 288 (230,371) |
| France | High income | 40+ threshold | 0.71 | 0.64 (0.52,0.69) | 0.64 (0.52,0.69) | 287 (230,370) | 287 (230,371) |
| France | High income | 65+ threshold | 0.58 | 0.65 (0.53,0.69) | 0.54 (0.39,0.68) | 286 (229,367) | 275 (221,369) |
| France | High income | Full sharing | 0.52 | 0.66 (0.54,0.71) | 0.20 (0.17,0.23) | 328 (259,425) | 135 (117,154) |
| Gabon | Upper middle income | Default | 0.02 | 0.54 (0.42,0.62) | 0.54 (0.42,0.62) | 131 (98,193) | 131 (98,193) |
| Gabon | Upper middle income | 2 dose threshold | 0.13 | 0.54 (0.42,0.61) | 0.54 (0.42,0.61) | 131 (97,193) | 131 (97,193) |
| Gabon | Upper middle income | 40+ threshold | 0.29 | 0.52 (0.40,0.60) | 0.52 (0.39,0.60) | 121 (80,185) | 121 (80,185) |
| Gabon | Upper middle income | 65+ threshold | 0.41 | 0.47 (0.32,0.59) | 0.44 (0.28,0.59) | 95 (59,142) | 93 (56,141) |
| Gabon | Upper middle income | Full sharing | 0.43 | 0.43 (0.30,0.56) | 0.16 (0.13,0.20) | 72 (48,100) | 29 (22,37) |
| United Kingdom | High income | Default | 1.00 | 0.36 (0.31,0.42) | 0.36 (0.31,0.42) | 248 (198,306) | 248 (198,306) |

| Country | Income bracket | Strategy | Proportion vaccinated | Proportion infected | | Mortalities per 100,000 | |
|---|---|---|---|---|---|---|---|
| | | | | unchanged | adapted | unchanged | adapted |
| United Kingdom | High income | 2 dose threshold | 0.94 | 0.36 (0.31,0.42) | 0.35 (0.30,0.42) | 248 (198,306) | 247 (197,306) |
| United Kingdom | High income | 40+ threshold | 0.70 | 0.63 (0.55,0.68) | 0.33 (0.27,0.42) | 277 (221,351) | 246 (196,303) |
| United Kingdom | High income | 65+ threshold | 0.59 | 0.75 (0.71,0.78) | 0.44 (0.34,0.55) | 319 (245,421) | 279 (214,358) |
| United Kingdom | High income | Full sharing | 0.52 | 0.80 (0.76,0.83) | 0.16 (0.15,0.18) | 547 (392,760) | 186 (156,223) |
| Georgia | Upper middle income | Default | 0.06 | 0.92 (0.90,0.94) | 0.92 (0.90,0.94) | 492 (402,580) | 492 (402,580) |
| Georgia | Upper middle income | 2 dose threshold | 0.17 | 0.92 (0.90,0.94) | 0.92 (0.90,0.94) | 492 (402,579) | 492 (402,579) |
| Georgia | Upper middle income | 40+ threshold | 0.35 | 0.92 (0.90,0.94) | 0.92 (0.90,0.94) | 487 (397,574) | 487 (397,574) |
| Georgia | Upper middle income | 65+ threshold | 0.49 | 0.92 (0.89,0.94) | 0.88 (0.85,0.91) | 473 (385,567) | 470 (383,567) |
| Georgia | Upper middle income | Full sharing | 0.51 | 0.92 (0.89,0.93) | 0.63 (0.59,0.67) | 447 (363,536) | 348 (297,404) |
| Ghana | Lower middle income | Default | 0.04 | 0.61 (0.46,0.73) | 0.61 (0.46,0.73) | 75 (53,96) | 75 (53,96) |
| Ghana | Lower middle income | 2 dose threshold | 0.14 | 0.61 (0.46,0.73) | 0.61 (0.46,0.73) | 75 (52,96) | 75 (52,96) |
| Ghana | Lower middle income | 40+ threshold | 0.30 | 0.58 (0.44,0.71) | 0.58 (0.45,0.71) | 70 (49,95) | 70 (49,95) |
| Ghana | Lower middle income | 65+ threshold | 0.42 | 0.49 (0.38,0.58) | 0.42 (0.33,0.50) | 62 (44,87) | 61 (43,84) |
| Ghana | Lower middle income | Full sharing | 0.43 | 0.45 (0.35,0.54) | 0.14 (0.10,0.17) | 55 (39,77) | 17 (12,21) |
| Guinea | Low income | Default | 0.04 | 0.72 (0.65,0.78) | 0.72 (0.65,0.78) | 77 (44,130) | 77 (44,130) |
| Guinea | Low income | 2 dose threshold | 0.14 | 0.72 (0.64,0.78) | 0.72 (0.64,0.78) | 77 (44,130) | 77 (44,130) |
| Guinea | Low income | 40+ threshold | 0.28 | 0.72 (0.64,0.78) | 0.72 (0.64,0.78) | 73 (42,123) | 73 (42,123) |
| Guinea | Low income | 65+ threshold | 0.38 | 0.67 (0.56,0.74) | 0.57 (0.39,0.69) | 58 (36,99) | 55 (33,99) |
| Guinea | Low income | Full sharing | 0.40 | 0.62 (0.46,0.71) | 0.21 (0.15,0.27) | 44 (29,72) | 19 (13,26) |
| Gambia | Low income | Default | 0.02 | 0.80 (0.72,0.86) | 0.80 (0.72,0.86) | 229 (150,342) | 229 (150,342) |
| Gambia | Low income | 2 dose threshold | 0.12 | 0.80 (0.72,0.86) | 0.80 (0.72,0.86) | 228 (150,342) | 228 (150,342) |
| Gambia | Low income | 40+ threshold | 0.26 | 0.80 (0.72,0.86) | 0.80 (0.72,0.86) | 212 (136,313) | 212 (136,312) |
| Gambia | Low income | 65+ threshold | 0.37 | 0.75 (0.67,0.81) | 0.58 (0.47,0.67) | 164 (107,234) | 150 (95,224) |
| Gambia | Low income | Full sharing | 0.40 | 0.69 (0.59,0.76) | 0.30 (0.25,0.35) | 135 (89,190) | 62 (43,86) |
| Guinea-Bissau | Low income | Default | 0.02 | 0.70 (0.61,0.77) | 0.70 (0.61,0.77) | 121 (69,177) | 121 (69,177) |
| Guinea-Bissau | Low income | 2 dose threshold | 0.12 | 0.70 (0.61,0.77) | 0.70 (0.61,0.77) | 121 (69,177) | 121 (69,177) |
| Guinea-Bissau | Low income | 40+ threshold | 0.27 | 0.70 (0.61,0.76) | 0.70 (0.61,0.76) | 113 (64,165) | 113 (64,165) |
| Guinea-Bissau | Low income | 65+ threshold | 0.39 | 0.62 (0.54,0.70) | 0.56 (0.46,0.67) | 89 (51,132) | 86 (51,127) |
| Guinea-Bissau | Low income | Full sharing | 0.41 | 0.58 (0.50,0.67) | 0.33 (0.26,0.41) | 76 (44,113) | 43 (27,57) |
| Equatorial Guinea | Upper middle income | Default | 0.16 | 0.62 (0.53,0.68) | 0.62 (0.53,0.68) | 139 (87,220) | 139 (87,220) |
| Equatorial Guinea | Upper middle income | 2 dose threshold | 0.25 | 0.62 (0.53,0.68) | 0.62 (0.53,0.68) | 139 (87,219) | 139 (87,219) |
| Equatorial Guinea | Upper middle income | 40+ threshold | 0.36 | 0.62 (0.52,0.68) | 0.61 (0.52,0.67) | 136 (85,214) | 136 (85,215) |
| Equatorial Guinea | Upper middle income | 65+ threshold | 0.42 | 0.60 (0.49,0.67) | 0.54 (0.42,0.66) | 122 (77,184) | 118 (75,182) |
| Equatorial Guinea | Upper middle income | Full sharing | 0.43 | 0.57 (0.45,0.65) | 0.30 (0.25,0.37) | 100 (64,148) | 54 (37,77) |
| Greece | High income | Default | 0.69 | 0.50 (0.21,0.76) | 0.50 (0.21,0.76) | 303 (159,454) | 303 (159,454) |

| Country | Income bracket | Strategy | Proportion vaccinated | Proportion infected | | Mortalities per 100,000 | |
|---|---|---|---|---|---|---|---|
| | | | | unchanged | adapted | unchanged | adapted |
| Greece | High income | 2 dose threshold | 0.75 | 0.50 (0.21,0.76) | 0.50 (0.21,0.76) | 303 (159,454) | 303 (159,454) |
| Greece | High income | 40+ threshold | 0.73 | 0.50 (0.20,0.76) | 0.50 (0.20,0.76) | 302 (157,453) | 302 (157,453) |
| Greece | High income | 65+ threshold | 0.60 | 0.50 (0.22,0.75) | 0.48 (0.18,0.75) | 288 (154,435) | 284 (147,435) |
| Greece | High income | Full sharing | 0.54 | 0.53 (0.25,0.78) | 0.06 (0.05,0.07) | 383 (191,575) | 58 (55,64) |
| Guatemala | Upper middle income | Default | 0.06 | 0.81 (0.77,0.86) | 0.81 (0.77,0.86) | 243 (134,369) | 243 (134,369) |
| Guatemala | Upper middle income | 2 dose threshold | 0.17 | 0.81 (0.77,0.86) | 0.81 (0.77,0.86) | 243 (134,368) | 243 (134,368) |
| Guatemala | Upper middle income | 40+ threshold | 0.32 | 0.81 (0.77,0.85) | 0.81 (0.77,0.85) | 235 (130,362) | 235 (130,362) |
| Guatemala | Upper middle income | 65+ threshold | 0.43 | 0.78 (0.73,0.83) | 0.71 (0.56,0.81) | 187 (111,286) | 180 (104,276) |
| Guatemala | Upper middle income | Full sharing | 0.45 | 0.76 (0.69,0.81) | 0.33 (0.27,0.40) | 147 (87,220) | 70 (46,93) |
| Guyana | Upper middle income | Default | 0.44 | 0.71 (0.59,0.81) | 0.71 (0.59,0.81) | 245 (146,380) | 245 (146,380) |
| Guyana | Upper middle income | 2 dose threshold | 0.51 | 0.71 (0.59,0.81) | 0.71 (0.59,0.81) | 245 (146,380) | 245 (146,380) |
| Guyana | Upper middle income | 40+ threshold | 0.52 | 0.71 (0.59,0.81) | 0.70 (0.50,0.81) | 242 (145,376) | 241 (142,376) |
| Guyana | Upper middle income | 65+ threshold | 0.47 | 0.72 (0.64,0.81) | 0.67 (0.41,0.81) | 223 (143,342) | 218 (128,342) |
| Guyana | Upper middle income | Full sharing | 0.48 | 0.71 (0.62,0.80) | 0.26 (0.20,0.34) | 186 (124,280) | 69 (48,91) |
| Honduras | Lower middle income | Default | 0.11 | 0.87 (0.84,0.88) | 0.87 (0.84,0.88) | 405 (234,617) | 405 (234,617) |
| Honduras | Lower middle income | 2 dose threshold | 0.23 | 0.87 (0.84,0.88) | 0.87 (0.84,0.88) | 405 (234,617) | 405 (234,617) |
| Honduras | Lower middle income | 40+ threshold | 0.35 | 0.87 (0.84,0.88) | 0.87 (0.84,0.88) | 394 (229,604) | 395 (229,604) |
| Honduras | Lower middle income | 65+ threshold | 0.45 | 0.85 (0.82,0.86) | 0.81 (0.74,0.86) | 334 (199,512) | 329 (198,509) |
| Honduras | Lower middle income | Full sharing | 0.47 | 0.83 (0.81,0.85) | 0.39 (0.34,0.45) | 269 (159,405) | 137 (95,183) |
| Croatia | High income | Default | 0.52 | 0.76 (0.68,0.81) | 0.76 (0.68,0.81) | 275 (204,363) | 275 (204,363) |
| Croatia | High income | 2 dose threshold | 0.60 | 0.76 (0.67,0.81) | 0.76 (0.67,0.81) | 275 (204,363) | 275 (204,363) |
| Croatia | High income | 40+ threshold | 0.69 | 0.76 (0.67,0.81) | 0.76 (0.67,0.81) | 274 (204,362) | 275 (204,362) |
| Croatia | High income | 65+ threshold | 0.58 | 0.76 (0.67,0.80) | 0.75 (0.61,0.81) | 270 (201,355) | 269 (200,352) |
| Croatia | High income | Full sharing | 0.53 | 0.76 (0.67,0.81) | 0.35 (0.29,0.40) | 283 (209,373) | 160 (129,198) |
| Hungary | High income | Default | 0.84 | 0.78 (0.64,0.84) | 0.78 (0.64,0.84) | 261 (214,316) | 261 (214,316) |
| Hungary | High income | 2 dose threshold | 0.87 | 0.78 (0.64,0.84) | 0.78 (0.64,0.84) | 260 (214,316) | 260 (214,316) |
| Hungary | High income | 40+ threshold | 0.74 | 0.78 (0.64,0.84) | 0.78 (0.64,0.84) | 260 (214,316) | 260 (214,316) |
| Hungary | High income | 65+ threshold | 0.59 | 0.78 (0.66,0.84) | 0.74 (0.58,0.85) | 261 (216,317) | 257 (212,314) |
| Hungary | High income | Full sharing | 0.53 | 0.79 (0.67,0.85) | 0.30 (0.23,0.35) | 288 (236,357) | 135 (120,148) |
| Indonesia | Upper middle income | Default | 0.16 | 0.87 (0.77,0.91) | 0.87 (0.77,0.91) | 165 (96,242) | 165 (96,242) |
| Indonesia | Upper middle income | 2 dose threshold | 0.27 | 0.87 (0.77,0.91) | 0.87 (0.77,0.91) | 165 (96,242) | 165 (96,242) |
| Indonesia | Upper middle income | 40+ threshold | 0.40 | 0.86 (0.76,0.91) | 0.86 (0.77,0.91) | 162 (94,235) | 162 (94,235) |
| Indonesia | Upper middle income | 65+ threshold | 0.48 | 0.84 (0.72,0.90) | 0.41 (0.31,0.53) | 140 (82,201) | 100 (61,144) |
| Indonesia | Upper middle income | Full sharing | 0.49 | 0.82 (0.66,0.89) | 0.14 (0.11,0.17) | 123 (72,177) | 32 (22,42) |
| India | Lower middle income | Default | 0.30 | 0.91 (0.82,0.94) | 0.91 (0.82,0.94) | 243 (179,306) | 243 (179,306) |

| Country | Income bracket | Strategy | Proportion vaccinated | Proportion infected | | Mortalities per 100,000 | |
|---|---|---|---|---|---|---|---|
| | | | | unchanged | adapted | unchanged | adapted |
| India | Lower middle income | 2 dose threshold | 0.39 | 0.91 (0.82,0.94) | 0.91 (0.82,0.94) | 243 (179,306) | 243 (179,306) |
| India | Lower middle income | 40+ threshold | 0.45 | 0.91 (0.82,0.94) | 0.91 (0.82,0.94) | 241 (178,304) | 241 (178,304) |
| India | Lower middle income | 65+ threshold | 0.48 | 0.90 (0.81,0.94) | 0.90 (0.81,0.94) | 225 (169,282) | 225 (169,281) |
| India | Lower middle income | Full sharing | 0.49 | 0.89 (0.80,0.93) | 0.31 (0.28,0.34) | 194 (147,242) | 78 (62,94) |
| Ireland | High income | Default | 0.81 | 0.39 (0.28,0.48) | 0.39 (0.28,0.48) | 145 (122,171) | 145 (122,171) |
| Ireland | High income | 2 dose threshold | 0.84 | 0.39 (0.28,0.48) | 0.39 (0.28,0.48) | 145 (122,171) | 145 (122,171) |
| Ireland | High income | 40+ threshold | 0.65 | 0.39 (0.28,0.49) | 0.40 (0.27,0.49) | 145 (123,170) | 145 (122,170) |
| Ireland | High income | 65+ threshold | 0.54 | 0.48 (0.35,0.58) | 0.26 (0.21,0.33) | 158 (136,179) | 135 (116,162) |
| Ireland | High income | Full sharing | 0.51 | 0.52 (0.38,0.61) | 0.09 (0.08,0.12) | 199 (164,230) | 54 (51,62) |
| Iran | Upper middle income | Default | 0.07 | 0.79 (0.76,0.83) | 0.79 (0.76,0.83) | 317 (209,457) | 317 (209,457) |
| Iran | Upper middle income | 2 dose threshold | 0.18 | 0.79 (0.76,0.83) | 0.79 (0.76,0.83) | 317 (209,456) | 317 (209,456) |
| Iran | Upper middle income | 40+ threshold | 0.36 | 0.79 (0.76,0.82) | 0.79 (0.76,0.82) | 310 (206,446) | 310 (207,447) |
| Iran | Upper middle income | 65+ threshold | 0.47 | 0.77 (0.72,0.81) | 0.74 (0.62,0.81) | 268 (179,383) | 264 (178,382) |
| Iran | Upper middle income | Full sharing | 0.49 | 0.76 (0.69,0.80) | 0.37 (0.31,0.45) | 220 (148,307) | 124 (94,159) |
| Iraq | Upper middle income | Default | 0.02 | 0.89 (0.83,0.94) | 0.89 (0.83,0.94) | 348 (188,534) | 348 (188,534) |
| Iraq | Upper middle income | 2 dose threshold | 0.13 | 0.89 (0.83,0.94) | 0.89 (0.83,0.94) | 348 (188,534) | 348 (188,534) |
| Iraq | Upper middle income | 40+ threshold | 0.29 | 0.89 (0.83,0.94) | 0.89 (0.83,0.94) | 338 (182,521) | 338 (182,522) |
| Iraq | Upper middle income | 65+ threshold | 0.41 | 0.88 (0.81,0.93) | 0.88 (0.79,0.93) | 292 (152,454) | 291 (152,453) |
| Iraq | Upper middle income | Full sharing | 0.43 | 0.86 (0.78,0.92) | 0.54 (0.48,0.60) | 220 (120,336) | 150 (99,209) |
| Iceland | High income | Default | 1.02 | 0.19 (0.15,0.23) | 0.19 (0.15,0.23) | 28 (24,33) | 28 (24,33) |
| Iceland | High income | 2 dose threshold | 0.92 | 0.19 (0.15,0.23) | 0.19 (0.15,0.23) | 28 (24,33) | 28 (24,33) |
| Iceland | High income | 40+ threshold | 0.66 | 0.21 (0.17,0.25) | 0.23 (0.16,0.26) | 28 (24,33) | 29 (25,34) |
| Iceland | High income | 65+ threshold | 0.56 | 0.28 (0.22,0.34) | 0.17 (0.13,0.21) | 31 (27,35) | 27 (23,32) |
| Iceland | High income | Full sharing | 0.52 | 0.31 (0.23,0.38) | 0.19 (0.16,0.23) | 36 (31,40) | 29 (24,34) |
| Israel | High income | Default | 0.88 | 0.45 (0.38,0.52) | 0.45 (0.38,0.52) | 57 (54,60) | 57 (54,60) |
| Israel | High income | 2 dose threshold | 0.80 | 0.52 (0.44,0.59) | 0.35 (0.29,0.40) | 61 (57,64) | 52 (47,58) |
| Israel | High income | 40+ threshold | 0.57 | 0.77 (0.72,0.80) | 0.40 (0.33,0.53) | 81 (74,89) | 55 (49,62) |
| Israel | High income | 65+ threshold | 0.52 | 0.78 (0.70,0.82) | 0.65 (0.53,0.71) | 95 (85,105) | 84 (65,100) |
| Israel | High income | Full sharing | 0.52 | 0.77 (0.70,0.82) | 0.16 (0.14,0.18) | 101 (91,112) | 45 (41,50) |
| Italy | High income | Default | 0.85 | 0.61 (0.45,0.69) | 0.61 (0.45,0.69) | 370 (298,456) | 370 (298,456) |
| Italy | High income | 2 dose threshold | 0.89 | 0.61 (0.45,0.69) | 0.61 (0.45,0.69) | 369 (298,456) | 369 (298,456) |
| Italy | High income | 40+ threshold | 0.77 | 0.60 (0.45,0.68) | 0.60 (0.45,0.69) | 368 (297,455) | 369 (297,455) |
| Italy | High income | 65+ threshold | 0.61 | 0.60 (0.45,0.68) | 0.53 (0.35,0.68) | 363 (292,443) | 353 (283,443) |
| Italy | High income | Full sharing | 0.54 | 0.62 (0.47,0.70) | 0.18 (0.14,0.22) | 425 (338,530) | 167 (150,178) |
| Jamaica | Upper middle income | Default | 0.08 | 0.55 (0.35,0.66) | 0.55 (0.35,0.66) | 210 (125,369) | 210 (125,369) |

| Country | Income bracket | Strategy | Proportion vaccinated | Proportion infected | | Mortalities per 100,000 | |
|---|---|---|---|---|---|---|---|
| | | | | unchanged | adapted | unchanged | adapted |
| Jamaica | Upper middle income | 2 dose threshold | 0.19 | 0.55 (0.35,0.66) | 0.55 (0.35,0.66) | 209 (125,369) | 209 (125,369) |
| Jamaica | Upper middle income | 40+ threshold | 0.37 | 0.54 (0.33,0.66) | 0.54 (0.33,0.66) | 203 (115,362) | 203 (115,362) |
| Jamaica | Upper middle income | 65+ threshold | 0.49 | 0.51 (0.29,0.65) | 0.46 (0.22,0.65) | 172 (86,315) | 166 (73,313) |
| Jamaica | Upper middle income | Full sharing | 0.50 | 0.48 (0.26,0.64) | 0.08 (0.06,0.11) | 130 (70,233) | 26 (20,33) |
| Japan | High income | Default | 0.37 | 0.36 (0.09,0.62) | 0.36 (0.09,0.62) | 184 (61,335) | 184 (61,335) |
| Japan | High income | 2 dose threshold | 0.48 | 0.36 (0.09,0.62) | 0.36 (0.09,0.62) | 184 (61,335) | 184 (61,335) |
| Japan | High income | 40+ threshold | 0.65 | 0.35 (0.08,0.62) | 0.36 (0.08,0.62) | 178 (56,329) | 179 (56,330) |
| Japan | High income | 65+ threshold | 0.56 | 0.30 (0.06,0.59) | 0.29 (0.06,0.59) | 129 (39,256) | 126 (37,254) |
| Japan | High income | Full sharing | 0.54 | 0.23 (0.05,0.52) | 0.02 (0.01,0.02) | 87 (31,180) | 12 (9,15) |
| Jordan | Upper middle income | Default | 0.37 | 0.82 (0.75,0.90) | 0.82 (0.75,0.90) | 207 (135,292) | 207 (135,292) |
| Jordan | Upper middle income | 2 dose threshold | 0.45 | 0.82 (0.75,0.90) | 0.82 (0.75,0.90) | 207 (135,292) | 207 (135,292) |
| Jordan | Upper middle income | 40+ threshold | 0.43 | 0.81 (0.75,0.90) | 0.82 (0.75,0.90) | 205 (134,290) | 205 (134,290) |
| Jordan | Upper middle income | 65+ threshold | 0.45 | 0.81 (0.75,0.90) | 0.81 (0.73,0.90) | 197 (130,279) | 197 (129,278) |
| Jordan | Upper middle income | Full sharing | 0.46 | 0.80 (0.74,0.89) | 0.43 (0.37,0.49) | 172 (114,240) | 82 (59,108) |
| Kazakhstan | Upper middle income | Default | 0.28 | 0.84 (0.79,0.89) | 0.84 (0.79,0.89) | 263 (144,437) | 263 (144,437) |
| Kazakhstan | Upper middle income | 2 dose threshold | 0.37 | 0.84 (0.79,0.89) | 0.84 (0.79,0.89) | 263 (144,437) | 263 (144,437) |
| Kazakhstan | Upper middle income | 40+ threshold | 0.45 | 0.84 (0.79,0.89) | 0.84 (0.79,0.89) | 258 (142,428) | 259 (142,428) |
| Kazakhstan | Upper middle income | 65+ threshold | 0.45 | 0.83 (0.78,0.87) | 0.73 (0.59,0.83) | 222 (123,359) | 211 (111,356) |
| Kazakhstan | Upper middle income | Full sharing | 0.47 | 0.82 (0.77,0.87) | 0.29 (0.21,0.36) | 190 (107,301) | 79 (54,104) |
| Kenya | Lower middle income | Default | 0.03 | 0.81 (0.75,0.84) | 0.81 (0.75,0.84) | 136 (82,206) | 136 (82,206) |
| Kenya | Lower middle income | 2 dose threshold | 0.13 | 0.81 (0.75,0.84) | 0.81 (0.75,0.84) | 136 (82,205) | 136 (82,205) |
| Kenya | Lower middle income | 40+ threshold | 0.29 | 0.81 (0.74,0.84) | 0.81 (0.74,0.84) | 133 (80,200) | 133 (80,200) |
| Kenya | Lower middle income | 65+ threshold | 0.41 | 0.79 (0.71,0.83) | 0.75 (0.60,0.83) | 115 (72,170) | 112 (68,165) |
| Kenya | Lower middle income | Full sharing | 0.43 | 0.76 (0.66,0.81) | 0.37 (0.28,0.44) | 92 (60,132) | 50 (37,65) |
| Kyrgyzstan | Lower middle income | Default | 0.04 | 0.88 (0.86,0.91) | 0.88 (0.86,0.91) | 317 (206,462) | 317 (206,462) |
| Kyrgyzstan | Lower middle income | 2 dose threshold | 0.15 | 0.88 (0.86,0.91) | 0.88 (0.86,0.91) | 316 (206,462) | 316 (206,462) |
| Kyrgyzstan | Lower middle income | 40+ threshold | 0.31 | 0.88 (0.86,0.91) | 0.88 (0.86,0.91) | 309 (201,448) | 309 (201,448) |
| Kyrgyzstan | Lower middle income | 65+ threshold | 0.43 | 0.87 (0.84,0.91) | 0.86 (0.81,0.91) | 272 (172,407) | 271 (170,406) |
| Kyrgyzstan | Lower middle income | Full sharing | 0.45 | 0.86 (0.83,0.90) | 0.57 (0.53,0.60) | 222 (144,327) | 163 (115,224) |
| South Korea | High income | Default | 0.00 | 0.28 (0.03,0.59) | 0.28 (0.03,0.59) | -152 (-2230,2382) | -152 (-2230,2382) |
| South Korea | High income | 2 dose threshold | 0.52 | 0.28 (0.03,0.59) | 0.28 (0.03,0.59) | 32 (-1585,2381) | 32 (-1585,2381) |
| South Korea | High income | 40+ threshold | 0.67 | 0.27 (0.03,0.59) | 0.28 (0.03,0.59) | 23 (-1534,2327) | 26 (-1545,2330) |
| South Korea | High income | 65+ threshold | 0.54 | 0.25 (0.02,0.57) | 0.24 (0.02,0.57) | -13 (-1050,1718) | -28 (-1009,1697) |
| South Korea | High income | Full sharing | 0.55 | 0.22 (0.02,0.55) | 0.01 (0.01,0.01) | -26 (-773,1361) | 6 (-13,44) |
| Kuwait | High income | Default | 0.70 | 0.77 (0.69,0.82) | 0.77 (0.69,0.82) | 84 (54,124) | 84 (54,124) |

| Country | Income bracket | Strategy | Proportion vaccinated | Proportion infected | | Mortalities per 100,000 | |
|---|---|---|---|---|---|---|---|
| | | | | unchanged | adapted | unchanged | adapted |
| Kuwait | High income | 2 dose threshold | 0.74 | 0.77 (0.69,0.82) | 0.77 (0.69,0.82) | 84 (54,124) | 84 (54,124) |
| Kuwait | High income | 40+ threshold | 0.58 | 0.77 (0.69,0.82) | 0.77 (0.70,0.82) | 84 (54,124) | 84 (55,124) |
| Kuwait | High income | 65+ threshold | 0.50 | 0.81 (0.76,0.85) | 0.60 (0.50,0.70) | 98 (62,149) | 79 (51,115) |
| Kuwait | High income | Full sharing | 0.52 | 0.80 (0.73,0.84) | 0.31 (0.23,0.38) | 90 (57,138) | 38 (29,48) |
| Liberia | Low income | Default | 0.02 | 0.74 (0.57,0.83) | 0.74 (0.57,0.83) | 123 (70,179) | 123 (70,179) |
| Liberia | Low income | 2 dose threshold | 0.12 | 0.74 (0.57,0.83) | 0.74 (0.57,0.83) | 123 (70,179) | 123 (70,179) |
| Liberia | Low income | 40+ threshold | 0.28 | 0.74 (0.56,0.83) | 0.74 (0.56,0.83) | 113 (65,166) | 114 (65,166) |
| Liberia | Low income | 65+ threshold | 0.40 | 0.69 (0.49,0.79) | 0.34 (0.24,0.43) | 66 (40,92) | 48 (31,67) |
| Liberia | Low income | Full sharing | 0.42 | 0.60 (0.40,0.72) | 0.18 (0.14,0.25) | 51 (32,69) | 23 (16,31) |
| Libya | Upper middle income | Default | 0.08 | 0.84 (0.77,0.89) | 0.84 (0.77,0.89) | 268 (164,364) | 268 (164,364) |
| Libya | Upper middle income | 2 dose threshold | 0.19 | 0.84 (0.77,0.89) | 0.84 (0.77,0.89) | 268 (164,364) | 268 (164,364) |
| Libya | Upper middle income | 40+ threshold | 0.36 | 0.83 (0.77,0.89) | 0.83 (0.77,0.89) | 255 (155,342) | 255 (155,342) |
| Libya | Upper middle income | 65+ threshold | 0.46 | 0.80 (0.75,0.84) | 0.70 (0.62,0.78) | 210 (133,284) | 202 (125,273) |
| Libya | Upper middle income | Full sharing | 0.48 | 0.78 (0.73,0.83) | 0.38 (0.34,0.43) | 173 (111,233) | 89 (62,115) |
| Sri Lanka | Lower middle income | Default | 0.19 | 0.62 (0.31,0.91) | 0.62 (0.31,0.91) | 34 (12,152) | 34 (12,152) |
| Sri Lanka | Lower middle income | 2 dose threshold | 0.30 | 0.62 (0.31,0.91) | 0.62 (0.31,0.91) | 42 (12,152) | 42 (12,152) |
| Sri Lanka | Lower middle income | 40+ threshold | 0.45 | 0.61 (0.30,0.91) | 0.62 (0.30,0.91) | 41 (11,142) | 41 (11,142) |
| Sri Lanka | Lower middle income | 65+ threshold | 0.50 | 0.57 (0.23,0.91) | 0.51 (0.12,0.91) | 34 (7,90) | 32 (6,88) |
| Sri Lanka | Lower middle income | Full sharing | 0.50 | 0.56 (0.22,0.91) | 0.02 (0.01,0.02) | 35 (7,96) | 2 (1,3) |
| Lithuania | High income | Default | 0.67 | 0.78 (0.68,0.82) | 0.78 (0.68,0.82) | 353 (246,512) | 353 (246,512) |
| Lithuania | High income | 2 dose threshold | 0.72 | 0.78 (0.68,0.82) | 0.78 (0.68,0.82) | 353 (246,512) | 353 (246,512) |
| Lithuania | High income | 40+ threshold | 0.73 | 0.77 (0.68,0.82) | 0.77 (0.68,0.82) | 352 (246,511) | 352 (246,511) |
| Lithuania | High income | 65+ threshold | 0.59 | 0.78 (0.70,0.83) | 0.70 (0.55,0.78) | 352 (247,508) | 345 (240,499) |
| Lithuania | High income | Full sharing | 0.53 | 0.80 (0.71,0.84) | 0.38 (0.28,0.48) | 387 (267,573) | 217 (171,289) |
| Luxembourg | High income | Default | 0.83 | 0.55 (0.45,0.62) | 0.55 (0.45,0.62) | 438 (374,510) | 438 (374,510) |
| Luxembourg | High income | 2 dose threshold | 0.87 | 0.55 (0.45,0.62) | 0.55 (0.45,0.62) | 438 (374,510) | 438 (374,510) |
| Luxembourg | High income | 40+ threshold | 0.69 | 0.55 (0.45,0.62) | 0.55 (0.45,0.62) | 437 (373,509) | 437 (373,509) |
| Luxembourg | High income | 65+ threshold | 0.57 | 0.56 (0.47,0.63) | 0.51 (0.41,0.60) | 436 (373,506) | 429 (368,497) |
| Luxembourg | High income | Full sharing | 0.53 | 0.57 (0.47,0.64) | 0.25 (0.21,0.29) | 451 (382,528) | 196 (178,214) |
| Latvia | High income | Default | 0.48 | 0.71 (0.63,0.78) | 0.71 (0.63,0.78) | 232 (140,391) | 232 (140,391) |
| Latvia | High income | 2 dose threshold | 0.56 | 0.71 (0.63,0.78) | 0.71 (0.63,0.78) | 232 (140,391) | 232 (140,391) |
| Latvia | High income | 40+ threshold | 0.66 | 0.71 (0.62,0.77) | 0.71 (0.62,0.77) | 231 (140,390) | 231 (140,390) |
| Latvia | High income | 65+ threshold | 0.56 | 0.71 (0.62,0.77) | 0.69 (0.55,0.76) | 224 (136,374) | 223 (135,372) |
| Latvia | High income | Full sharing | 0.52 | 0.71 (0.63,0.78) | 0.23 (0.17,0.30) | 232 (140,391) | 90 (64,117) |
| Morocco | Lower middle income | Default | 0.40 | 0.79 (0.70,0.87) | 0.79 (0.70,0.87) | 249 (142,358) | 249 (142,358) |

| **Country** | **Income bracket** | **Strategy** | **Proportion vaccinated** | **Proportion infected** | | **Mortalities per 100,000** | |
|---|---|---|---|---|---|---|---|
| | | | | unchanged | adapted | unchanged | adapted |
| Morocco | Lower middle income | 2 dose threshold | 0.48 | 0.79 (0.70,0.87) | 0.79 (0.70,0.87) | 249 (142,358) | 249 (142,358) |
| Morocco | Lower middle income | 40+ threshold | 0.53 | 0.78 (0.70,0.86) | 0.78 (0.70,0.86) | 248 (141,357) | 248 (141,357) |
| Morocco | Lower middle income | 65+ threshold | 0.51 | 0.79 (0.71,0.87) | 0.63 (0.50,0.77) | 248 (141,352) | 234 (134,348) |
| Morocco | Lower middle income | Full sharing | 0.48 | 0.80 (0.72,0.88) | 0.40 (0.33,0.48) | 240 (139,347) | 148 (100,202) |
| Moldova | Lower middle income | Default | 0.16 | 0.77 (0.70,0.82) | 0.77 (0.70,0.82) | 282 (137,541) | 282 (137,541) |
| Moldova | Lower middle income | 2 dose threshold | 0.27 | 0.77 (0.70,0.82) | 0.77 (0.70,0.82) | 284 (137,541) | 284 (137,541) |
| Moldova | Lower middle income | 40+ threshold | 0.46 | 0.77 (0.70,0.82) | 0.77 (0.70,0.82) | 282 (137,535) | 282 (137,536) |
| Moldova | Lower middle income | 65+ threshold | 0.52 | 0.76 (0.68,0.82) | 0.73 (0.64,0.81) | 270 (132,503) | 268 (132,503) |
| Moldova | Lower middle income | Full sharing | 0.53 | 0.75 (0.66,0.81) | 0.43 (0.33,0.52) | 235 (119,425) | 135 (94,181) |
| Madagascar | Low income | Default | 0.01 | 0.86 (0.78,0.91) | 0.86 (0.78,0.91) | 111 (63,179) | 111 (63,179) |
| Madagascar | Low income | 2 dose threshold | 0.11 | 0.86 (0.78,0.91) | 0.86 (0.78,0.91) | 111 (63,179) | 111 (63,179) |
| Madagascar | Low income | 40+ threshold | 0.27 | 0.86 (0.78,0.91) | 0.86 (0.78,0.91) | 109 (62,175) | 109 (62,175) |
| Madagascar | Low income | 65+ threshold | 0.40 | 0.86 (0.77,0.90) | 0.86 (0.77,0.90) | 95 (54,152) | 95 (54,152) |
| Madagascar | Low income | Full sharing | 0.42 | 0.84 (0.74,0.89) | 0.30 (0.23,0.38) | 72 (42,112) | 30 (21,38) |
| Maldives | Upper middle income | Default | 0.90 | 0.70 (0.51,0.82) | 0.70 (0.51,0.82) | 73 (48,109) | 73 (48,109) |
| Maldives | Upper middle income | 2 dose threshold | 0.91 | 0.70 (0.51,0.82) | 0.70 (0.51,0.82) | 73 (48,109) | 73 (48,109) |
| Maldives | Upper middle income | 40+ threshold | 0.48 | 0.86 (0.75,0.92) | 0.38 (0.32,0.43) | 86 (55,125) | 56 (38,77) |
| Maldives | Upper middle income | 65+ threshold | 0.51 | 0.94 (0.89,0.97) | 0.42 (0.32,0.49) | 130 (81,202) | 61 (40,87) |
| Maldives | Upper middle income | Full sharing | 0.52 | 0.94 (0.87,0.97) | 0.19 (0.16,0.24) | 116 (72,180) | 25 (17,34) |
| Mexico | Upper middle income | Default | 0.37 | 0.80 (0.76,0.84) | 0.80 (0.76,0.84) | 450 (319,591) | 450 (319,591) |
| Mexico | Upper middle income | 2 dose threshold | 0.46 | 0.80 (0.76,0.84) | 0.80 (0.76,0.84) | 450 (319,591) | 450 (319,591) |
| Mexico | Upper middle income | 40+ threshold | 0.50 | 0.79 (0.75,0.84) | 0.79 (0.75,0.83) | 448 (317,590) | 448 (317,590) |
| Mexico | Upper middle income | 65+ threshold | 0.49 | 0.79 (0.75,0.83) | 0.72 (0.67,0.80) | 441 (313,580) | 434 (301,579) |
| Mexico | Upper middle income | Full sharing | 0.49 | 0.79 (0.74,0.83) | 0.47 (0.44,0.50) | 418 (299,543) | 262 (203,311) |
| Republic of Macedonia | Upper middle income | Default | 0.52 | 0.86 (0.82,0.88) | 0.86 (0.82,0.88) | 460 (316,637) | 460 (316,637) |
| Republic of Macedonia | Upper middle income | 2 dose threshold | 0.60 | 0.85 (0.82,0.88) | 0.85 (0.82,0.88) | 460 (316,637) | 460 (316,637) |
| Republic of Macedonia | Upper middle income | 40+ threshold | 0.65 | 0.85 (0.82,0.88) | 0.85 (0.82,0.88) | 457 (315,635) | 457 (315,635) |
| Republic of Macedonia | Upper middle income | 65+ threshold | 0.52 | 0.85 (0.82,0.88) | 0.83 (0.77,0.88) | 443 (307,620) | 441 (307,619) |
| Republic of Macedonia | Upper middle income | Full sharing | 0.53 | 0.85 (0.81,0.88) | 0.55 (0.49,0.59) | 401 (277,558) | 280 (205,368) |
| Mali | Low income | Default | 0.01 | 0.74 (0.64,0.80) | 0.74 (0.64,0.80) | 108 (65,159) | 108 (65,159) |
| Mali | Low income | 2 dose threshold | 0.10 | 0.74 (0.64,0.80) | 0.74 (0.64,0.80) | 108 (65,159) | 108 (65,159) |
| Mali | Low income | 40+ threshold | 0.25 | 0.74 (0.64,0.80) | 0.74 (0.64,0.80) | 104 (63,155) | 105 (63,155) |
| Mali | Low income | 65+ threshold | 0.36 | 0.72 (0.60,0.79) | 0.71 (0.56,0.79) | 90 (54,137) | 89 (54,132) |
| Mali | Low income | Full sharing | 0.38 | 0.68 (0.55,0.77) | 0.34 (0.27,0.42) | 70 (44,103) | 41 (29,51) |
| Malta | High income | Default | 1.06 | 0.39 (0.28,0.53) | 0.39 (0.28,0.53) | 216 (169,281) | 216 (169,281) |

| **Country** | **Income bracket** | **Strategy** | **Proportion vaccinated** | **Proportion infected** | | **Mortalities per 100,000** | |
|---|---|---|---|---|---|---|---|
| | | | | unchanged | adapted | unchanged | adapted |
| Malta | High income | 2 dose threshold | 0.99 | 0.39 (0.29,0.53) | 0.39 (0.28,0.53) | 216 (169,281) | 216 (168,281) |
| Malta | High income | 40+ threshold | 0.73 | 0.42 (0.33,0.54) | 0.39 (0.30,0.53) | 217 (172,282) | 216 (169,281) |
| Malta | High income | 65+ threshold | 0.60 | 0.50 (0.46,0.56) | 0.41 (0.32,0.52) | 234 (189,301) | 222 (174,289) |
| Malta | High income | Full sharing | 0.53 | 0.54 (0.50,0.60) | 0.19 (0.16,0.25) | 311 (242,409) | 129 (105,167) |
| Myanmar | Lower middle income | Default | 0.07 | 0.83 (0.56,0.96) | 0.83 (0.56,0.96) | 203 (128,305) | 203 (128,305) |
| Myanmar | Lower middle income | 2 dose threshold | 0.19 | 0.83 (0.56,0.96) | 0.83 (0.56,0.96) | 203 (127,305) | 203 (127,305) |
| Myanmar | Lower middle income | 40+ threshold | 0.36 | 0.82 (0.55,0.96) | 0.82 (0.55,0.96) | 188 (118,283) | 188 (118,283) |
| Myanmar | Lower middle income | 65+ threshold | 0.48 | 0.77 (0.45,0.94) | 0.21 (0.15,0.29) | 125 (79,189) | 64 (45,88) |
| Myanmar | Lower middle income | Full sharing | 0.50 | 0.72 (0.37,0.92) | 0.10 (0.08,0.12) | 105 (66,161) | 28 (20,37) |
| Montenegro | Upper middle income | Default | 0.34 | 0.88 (0.88,0.89) | 0.88 (0.88,0.89) | 375 (276,475) | 375 (276,475) |
| Montenegro | Upper middle income | 2 dose threshold | 0.44 | 0.88 (0.88,0.89) | 0.88 (0.88,0.89) | 375 (276,475) | 375 (276,475) |
| Montenegro | Upper middle income | 40+ threshold | 0.57 | 0.88 (0.87,0.89) | 0.88 (0.87,0.89) | 374 (275,474) | 374 (275,474) |
| Montenegro | Upper middle income | 65+ threshold | 0.53 | 0.88 (0.87,0.89) | 0.85 (0.82,0.87) | 368 (271,465) | 366 (269,463) |
| Montenegro | Upper middle income | Full sharing | 0.52 | 0.88 (0.87,0.89) | 0.52 (0.49,0.56) | 343 (253,434) | 216 (165,268) |
| Mozambique | Low income | Default | 0.02 | 0.81 (0.68,0.88) | 0.81 (0.68,0.88) | 113 (60,177) | 113 (60,177) |
| Mozambique | Low income | 2 dose threshold | 0.11 | 0.81 (0.68,0.88) | 0.81 (0.68,0.88) | 113 (60,177) | 113 (60,177) |
| Mozambique | Low income | 40+ threshold | 0.26 | 0.81 (0.68,0.88) | 0.81 (0.68,0.88) | 103 (55,162) | 103 (55,162) |
| Mozambique | Low income | 65+ threshold | 0.38 | 0.72 (0.56,0.81) | 0.42 (0.27,0.54) | 66 (39,95) | 56 (34,76) |
| Mozambique | Low income | Full sharing | 0.40 | 0.60 (0.41,0.73) | 0.11 (0.07,0.15) | 55 (34,77) | 12 (8,14) |
| Mauritania | Lower middle income | Default | 0.05 | 0.76 (0.66,0.83) | 0.76 (0.66,0.83) | 157 (97,233) | 157 (97,233) |
| Mauritania | Lower middle income | 2 dose threshold | 0.15 | 0.76 (0.66,0.83) | 0.76 (0.66,0.83) | 157 (97,233) | 157 (97,233) |
| Mauritania | Lower middle income | 40+ threshold | 0.28 | 0.75 (0.66,0.83) | 0.75 (0.66,0.83) | 145 (88,219) | 146 (88,219) |
| Mauritania | Lower middle income | 65+ threshold | 0.40 | 0.68 (0.59,0.77) | 0.52 (0.41,0.63) | 106 (63,160) | 97 (57,150) |
| Mauritania | Lower middle income | Full sharing | 0.42 | 0.62 (0.50,0.73) | 0.32 (0.25,0.38) | 88 (54,130) | 53 (37,73) |
| Mauritius | High income | Default | 0.63 | 0.27 (0.13,0.44) | 0.27 (0.13,0.44) | 30 (20,44) | 30 (20,44) |
| Mauritius | High income | 2 dose threshold | 0.70 | 0.27 (0.13,0.44) | 0.27 (0.13,0.44) | 30 (20,44) | 30 (20,44) |
| Mauritius | High income | 40+ threshold | 0.59 | 0.26 (0.12,0.43) | 0.26 (0.12,0.43) | 29 (19,43) | 29 (19,43) |
| Mauritius | High income | 65+ threshold | 0.55 | 0.27 (0.12,0.45) | 0.07 (0.04,0.13) | 32 (20,45) | 15 (8,29) |
| Mauritius | High income | Full sharing | 0.53 | 0.29 (0.12,0.49) | 0.05 (0.04,0.07) | 38 (21,56) | 13 (9,19) |
| Malawi | Low income | Default | 0.03 | 0.83 (0.69,0.89) | 0.83 (0.69,0.89) | 67 (38,106) | 67 (38,106) |
| Malawi | Low income | 2 dose threshold | 0.13 | 0.83 (0.69,0.89) | 0.83 (0.69,0.89) | 67 (38,105) | 67 (38,105) |
| Malawi | Low income | 40+ threshold | 0.28 | 0.82 (0.68,0.89) | 0.82 (0.68,0.89) | 64 (37,102) | 64 (37,102) |
| Malawi | Low income | 65+ threshold | 0.39 | 0.75 (0.62,0.83) | 0.63 (0.55,0.70) | 58 (34,96) | 56 (33,95) |
| Malawi | Low income | Full sharing | 0.41 | 0.71 (0.59,0.78) | 0.18 (0.13,0.23) | 51 (30,86) | 14 (10,19) |
| Malaysia | Upper middle income | Default | 0.29 | 0.75 (0.56,0.92) | 0.75 (0.56,0.92) | 100 (59,173) | 100 (59,173) |

| Country | Income bracket | Strategy | Proportion vaccinated | Proportion infected | | Mortalities per 100,000 | |
|---|---|---|---|---|---|---|---|
| | | | | unchanged | adapted | unchanged | adapted |
| Malaysia | Upper middle income | 2 dose threshold | 0.39 | 0.75 (0.56,0.92) | 0.75 (0.56,0.92) | 100 (59,172) | 100 (59,172) |
| Malaysia | Upper middle income | 40+ threshold | 0.44 | 0.75 (0.55,0.92) | 0.75 (0.55,0.92) | 97 (55,170) | 97 (55,171) |
| Malaysia | Upper middle income | 65+ threshold | 0.49 | 0.72 (0.51,0.91) | 0.61 (0.16,0.91) | 75 (34,145) | 71 (21,144) |
| Malaysia | Upper middle income | Full sharing | 0.50 | 0.69 (0.47,0.89) | 0.03 (0.02,0.03) | 54 (27,102) | 3 (2,4) |
| Namibia | Upper middle income | Default | 0.07 | 0.92 (0.89,0.94) | 0.92 (0.89,0.94) | 182 (106,283) | 182 (106,283) |
| Namibia | Upper middle income | 2 dose threshold | 0.17 | 0.92 (0.89,0.94) | 0.92 (0.89,0.94) | 182 (106,282) | 182 (106,282) |
| Namibia | Upper middle income | 40+ threshold | 0.32 | 0.92 (0.89,0.94) | 0.92 (0.89,0.94) | 173 (101,270) | 173 (101,270) |
| Namibia | Upper middle income | 65+ threshold | 0.41 | 0.91 (0.88,0.93) | 0.90 (0.86,0.93) | 127 (77,202) | 126 (75,201) |
| Namibia | Upper middle income | Full sharing | 0.44 | 0.89 (0.85,0.91) | 0.26 (0.22,0.31) | 90 (58,137) | 31 (23,42) |
| Niger | Low income | Default | 0.01 | 0.47 (0.39,0.55) | 0.47 (0.39,0.55) | 43 (27,60) | 43 (27,60) |
| Niger | Low income | 2 dose threshold | 0.10 | 0.47 (0.38,0.54) | 0.47 (0.38,0.54) | 43 (27,60) | 43 (27,60) |
| Niger | Low income | 40+ threshold | 0.24 | 0.46 (0.38,0.53) | 0.46 (0.37,0.53) | 42 (26,59) | 42 (26,59) |
| Niger | Low income | 65+ threshold | 0.34 | 0.45 (0.35,0.53) | 0.44 (0.35,0.52) | 40 (25,57) | 40 (24,57) |
| Niger | Low income | Full sharing | 0.36 | 0.44 (0.34,0.52) | 0.23 (0.17,0.29) | 35 (22,49) | 19 (14,24) |
| Nigeria | Lower middle income | Default | 0.02 | 0.62 (0.52,0.72) | 0.62 (0.52,0.72) | 59 (30,86) | 59 (30,86) |
| Nigeria | Lower middle income | 2 dose threshold | 0.11 | 0.61 (0.52,0.71) | 0.61 (0.52,0.71) | 59 (30,86) | 59 (30,86) |
| Nigeria | Lower middle income | 40+ threshold | 0.26 | 0.59 (0.48,0.70) | 0.59 (0.48,0.70) | 57 (30,84) | 57 (30,84) |
| Nigeria | Lower middle income | 65+ threshold | 0.38 | 0.58 (0.47,0.70) | 0.58 (0.47,0.70) | 56 (29,82) | 56 (29,82) |
| Nigeria | Lower middle income | Full sharing | 0.40 | 0.58 (0.46,0.69) | 0.29 (0.22,0.37) | 49 (26,72) | 23 (14,31) |
| Nicaragua | Lower middle income | Default | 0.05 | 0.80 (0.76,0.83) | 0.80 (0.76,0.83) | 281 (227,353) | 281 (227,353) |
| Nicaragua | Lower middle income | 2 dose threshold | 0.16 | 0.80 (0.76,0.83) | 0.80 (0.76,0.83) | 281 (227,353) | 281 (227,353) |
| Nicaragua | Lower middle income | 40+ threshold | 0.33 | 0.80 (0.76,0.83) | 0.80 (0.76,0.83) | 270 (219,338) | 270 (219,340) |
| Nicaragua | Lower middle income | 65+ threshold | 0.45 | 0.77 (0.73,0.80) | 0.69 (0.58,0.76) | 220 (176,275) | 211 (161,265) |
| Nicaragua | Lower middle income | Full sharing | 0.47 | 0.74 (0.68,0.78) | 0.35 (0.32,0.38) | 183 (148,229) | 95 (79,114) |
| Netherlands | High income | Default | 0.92 | 0.59 (0.44,0.66) | 0.59 (0.44,0.66) | 451 (288,644) | 451 (288,644) |
| Netherlands | High income | 2 dose threshold | 0.94 | 0.59 (0.44,0.66) | 0.59 (0.44,0.66) | 451 (288,644) | 451 (288,644) |
| Netherlands | High income | 40+ threshold | 0.72 | 0.59 (0.44,0.65) | 0.59 (0.44,0.66) | 449 (287,641) | 449 (287,642) |
| Netherlands | High income | 65+ threshold | 0.58 | 0.59 (0.44,0.65) | 0.49 (0.32,0.68) | 442 (290,624) | 420 (269,633) |
| Netherlands | High income | Full sharing | 0.53 | 0.60 (0.44,0.68) | 0.17 (0.14,0.20) | 511 (325,739) | 171 (126,212) |
| Norway | High income | Default | 0.77 | 0.64 (0.41,0.77) | 0.64 (0.41,0.77) | 15 (11,27) | 15 (11,27) |
| Norway | High income | 2 dose threshold | 0.83 | 0.64 (0.41,0.77) | 0.64 (0.41,0.77) | 15 (11,27) | 15 (11,27) |
| Norway | High income | 40+ threshold | 0.66 | 0.63 (0.40,0.77) | 0.63 (0.41,0.77) | 15 (11,27) | 15 (11,27) |
| Norway | High income | 65+ threshold | 0.57 | 0.64 (0.44,0.76) | 0.59 (0.33,0.75) | 15 (11,26) | 15 (10,25) |
| Norway | High income | Full sharing | 0.52 | 0.66 (0.47,0.78) | 0.24 (0.12,0.39) | 18 (11,33) | 9 (9,10) |
| Nepal | Lower middle income | Default | 0.13 | 0.93 (0.88,0.96) | 0.93 (0.88,0.96) | 234 (143,336) | 234 (143,336) |

| Country | Income bracket | Strategy | Proportion vaccinated | Proportion infected | | Mortalities per 100,000 | |
|---|---|---|---|---|---|---|---|
| | | | | unchanged | adapted | unchanged | adapted |
| Nepal | Lower middle income | 2 dose threshold | 0.24 | 0.93 (0.88,0.96) | 0.93 (0.88,0.96) | 234 (143,335) | 234 (143,335) |
| Nepal | Lower middle income | 40+ threshold | 0.39 | 0.93 (0.88,0.96) | 0.93 (0.88,0.96) | 233 (142,333) | 233 (142,334) |
| Nepal | Lower middle income | 65+ threshold | 0.48 | 0.94 (0.88,0.97) | 0.58 (0.49,0.69) | 243 (151,350) | 180 (113,252) |
| Nepal | Lower middle income | Full sharing | 0.48 | 0.93 (0.88,0.97) | 0.32 (0.28,0.36) | 231 (143,333) | 91 (62,117) |
| New Zealand | High income | Default | 0.21 | 0.01 (0.01,0.01) | 0.01 (0.01,0.01) | 5 (4,6) | 5 (4,6) |
| New Zealand | High income | 2 dose threshold | 0.32 | 0.01 (0.01,0.01) | 0.01 (0.01,0.01) | 5 (4,6) | 5 (4,6) |
| New Zealand | High income | 40+ threshold | 0.49 | 0.01 (0.01,0.01) | 0.01 (0.01,0.01) | 5 (4,5) | 5 (4,5) |
| New Zealand | High income | 65+ threshold | 0.51 | 0.01 (0.01,0.01) | 0.01 (0.00,0.01) | 4 (3,4) | 4 (3,4) |
| New Zealand | High income | Full sharing | 0.51 | 0.01 (0.01,0.01) | 0.02 (0.01,0.04) | 3 (3,4) | 11 (4,18) |
| Oman | High income | Default | 0.30 | 0.77 (0.66,0.83) | 0.77 (0.66,0.83) | 203 (125,296) | 203 (125,296) |
| Oman | High income | 2 dose threshold | 0.40 | 0.77 (0.66,0.83) | 0.77 (0.66,0.83) | 203 (125,295) | 203 (125,295) |
| Oman | High income | 40+ threshold | 0.39 | 0.77 (0.65,0.82) | 0.77 (0.65,0.82) | 200 (123,292) | 200 (123,292) |
| Oman | High income | 65+ threshold | 0.49 | 0.76 (0.63,0.82) | 0.48 (0.38,0.58) | 187 (117,278) | 126 (82,182) |
| Oman | High income | Full sharing | 0.51 | 0.72 (0.57,0.80) | 0.23 (0.20,0.26) | 155 (97,228) | 64 (45,83) |
| Pakistan | Lower middle income | Default | 0.08 | 0.86 (0.83,0.89) | 0.86 (0.83,0.89) | 142 (95,207) | 142 (95,207) |
| Pakistan | Lower middle income | 2 dose threshold | 0.19 | 0.86 (0.83,0.89) | 0.86 (0.83,0.89) | 142 (95,207) | 142 (95,207) |
| Pakistan | Lower middle income | 40+ threshold | 0.33 | 0.86 (0.82,0.89) | 0.86 (0.82,0.89) | 140 (94,202) | 141 (94,202) |
| Pakistan | Lower middle income | 65+ threshold | 0.42 | 0.85 (0.80,0.89) | 0.84 (0.78,0.89) | 131 (89,180) | 131 (89,179) |
| Pakistan | Lower middle income | Full sharing | 0.45 | 0.84 (0.78,0.88) | 0.48 (0.41,0.55) | 112 (78,153) | 81 (61,103) |
| Panama | High income | Default | 0.94 | 0.67 (0.62,0.71) | 0.67 (0.62,0.71) | 250 (178,387) | 250 (178,387) |
| Panama | High income | 2 dose threshold | 0.89 | 0.67 (0.62,0.71) | 0.67 (0.62,0.71) | 250 (178,387) | 250 (178,387) |
| Panama | High income | 40+ threshold | 0.54 | 0.67 (0.62,0.71) | 0.67 (0.63,0.71) | 249 (177,387) | 249 (177,387) |
| Panama | High income | 65+ threshold | 0.50 | 0.68 (0.63,0.72) | 0.63 (0.57,0.70) | 247 (176,383) | 244 (173,378) |
| Panama | High income | Full sharing | 0.49 | 0.68 (0.63,0.73) | 0.41 (0.36,0.46) | 229 (166,350) | 139 (113,175) |
| Peru | Upper middle income | Default | 0.21 | 0.82 (0.79,0.86) | 0.82 (0.79,0.86) | 999 (607,1547) | 999 (607,1547) |
| Peru | Upper middle income | 2 dose threshold | 0.31 | 0.82 (0.79,0.86) | 0.82 (0.79,0.86) | 999 (607,1547) | 999 (607,1547) |
| Peru | Upper middle income | 40+ threshold | 0.44 | 0.82 (0.79,0.86) | 0.82 (0.79,0.86) | 990 (600,1527) | 991 (600,1528) |
| Peru | Upper middle income | 65+ threshold | 0.49 | 0.81 (0.76,0.84) | 0.78 (0.71,0.84) | 938 (564,1408) | 929 (559,1378) |
| Peru | Upper middle income | Full sharing | 0.49 | 0.80 (0.75,0.84) | 0.47 (0.40,0.54) | 817 (502,1210) | 410 (332,484) |
| Philippines | Lower middle income | Default | 0.11 | 0.71 (0.55,0.80) | 0.71 (0.55,0.80) | 193 (80,411) | 193 (80,411) |
| Philippines | Lower middle income | 2 dose threshold | 0.22 | 0.71 (0.55,0.80) | 0.71 (0.55,0.80) | 193 (80,411) | 193 (80,411) |
| Philippines | Lower middle income | 40+ threshold | 0.37 | 0.70 (0.52,0.80) | 0.70 (0.52,0.80) | 186 (78,401) | 186 (78,401) |
| Philippines | Lower middle income | 65+ threshold | 0.45 | 0.67 (0.43,0.80) | 0.64 (0.35,0.80) | 152 (65,340) | 150 (62,339) |
| Philippines | Lower middle income | Full sharing | 0.47 | 0.64 (0.39,0.78) | 0.12 (0.09,0.15) | 105 (47,230) | 25 (16,33) |
| Poland | High income | Default | 0.66 | 0.79 (0.69,0.85) | 0.79 (0.69,0.85) | 304 (167,543) | 304 (167,543) |

| Country | Income bracket | Strategy | Proportion vaccinated | Proportion infected | | Mortalities per 100,000 | |
|---|---|---|---|---|---|---|---|
| | | | | unchanged | adapted | unchanged | adapted |
| Poland | High income | 2 dose threshold | 0.72 | 0.79 (0.69,0.85) | 0.79 (0.69,0.85) | 304 (167,543) | 304 (167,543) |
| Poland | High income | 40+ threshold | 0.70 | 0.79 (0.69,0.85) | 0.79 (0.69,0.85) | 304 (167,542) | 304 (167,542) |
| Poland | High income | 65+ threshold | 0.58 | 0.78 (0.68,0.85) | 0.78 (0.66,0.85) | 302 (167,536) | 301 (167,536) |
| Poland | High income | Full sharing | 0.53 | 0.78 (0.67,0.85) | 0.29 (0.22,0.34) | 322 (174,580) | 178 (119,254) |
| Puerto Rico | High income | Default | 0.00 | 0.51 (0.38,0.59) | 0.51 (0.38,0.59) | 442 (207,949) | 442 (207,949) |
| Puerto Rico | High income | 2 dose threshold | 0.13 | 0.51 (0.38,0.59) | 0.51 (0.38,0.59) | 441 (207,948) | 441 (207,948) |
| Puerto Rico | High income | 40+ threshold | 0.32 | 0.51 (0.37,0.59) | 0.51 (0.37,0.59) | 428 (193,929) | 428 (193,931) |
| Puerto Rico | High income | 65+ threshold | 0.50 | 0.48 (0.33,0.58) | 0.46 (0.30,0.58) | 355 (153,769) | 350 (148,762) |
| Puerto Rico | High income | Full sharing | 0.54 | 0.45 (0.30,0.57) | 0.17 (0.14,0.21) | 298 (135,617) | 115 (78,171) |
| Portugal | High income | Default | 0.81 | 0.46 (0.40,0.56) | 0.46 (0.40,0.56) | 817 (-7293,5157) | 817 (-7293,5157) |
| Portugal | High income | 2 dose threshold | 0.86 | 0.43 (0.36,0.49) | 0.43 (0.36,0.49) | 1181 (-4464,5157) | 1181 (-4464,5157) |
| Portugal | High income | 40+ threshold | 0.75 | 0.43 (0.36,0.49) | 0.43 (0.36,0.49) | 1180 (-4460,5151) | 1180 (-4462,5147) |
| Portugal | High income | 65+ threshold | 0.61 | 0.46 (0.37,0.51) | 0.42 (0.34,0.48) | 1183 (-4471,5180) | 1170 (-4412,5085) |
| Portugal | High income | Full sharing | 0.54 | 0.48 (0.39,0.54) | 0.14 (0.12,0.16) | 1306 (-5033,5787) | 195 (-88,434) |
| Paraguay | Upper middle income | Default | 0.16 | 0.88 (0.83,0.92) | 0.88 (0.83,0.92) | 306 (198,500) | 306 (198,500) |
| Paraguay | Upper middle income | 2 dose threshold | 0.26 | 0.88 (0.83,0.92) | 0.88 (0.83,0.92) | 306 (197,500) | 306 (197,500) |
| Paraguay | Upper middle income | 40+ threshold | 0.39 | 0.88 (0.83,0.92) | 0.88 (0.83,0.92) | 301 (194,492) | 301 (194,492) |
| Paraguay | Upper middle income | 65+ threshold | 0.46 | 0.88 (0.82,0.91) | 0.88 (0.81,0.91) | 259 (165,426) | 258 (165,425) |
| Paraguay | Upper middle income | Full sharing | 0.48 | 0.86 (0.79,0.90) | 0.24 (0.20,0.27) | 192 (125,310) | 51 (41,63) |
| Palestine | Lower middle income | Default | 0.15 | 0.83 (0.79,0.87) | 0.83 (0.79,0.87) | 218 (109,418) | 218 (109,418) |
| Palestine | Lower middle income | 2 dose threshold | 0.24 | 0.83 (0.79,0.87) | 0.83 (0.79,0.87) | 218 (109,418) | 218 (109,418) |
| Palestine | Lower middle income | 40+ threshold | 0.35 | 0.82 (0.78,0.87) | 0.82 (0.78,0.87) | 214 (107,406) | 214 (107,406) |
| Palestine | Lower middle income | 65+ threshold | 0.41 | 0.81 (0.76,0.87) | 0.80 (0.73,0.86) | 197 (99,384) | 196 (97,382) |
| Palestine | Lower middle income | Full sharing | 0.43 | 0.80 (0.73,0.86) | 0.46 (0.40,0.50) | 151 (81,285) | 73 (51,105) |
| Qatar | High income | Default | 0.94 | 0.77 (0.68,0.83) | 0.77 (0.68,0.83) | 13 (21,76) | 13 (21,76) |
| Qatar | High income | 2 dose threshold | 0.96 | 0.77 (0.68,0.83) | 0.77 (0.68,0.83) | 24 (21,76) | 24 (21,76) |
| Qatar | High income | 40+ threshold | 0.52 | 0.79 (0.72,0.84) | 0.76 (0.67,0.84) | 25 (21,80) | 24 (21,75) |
| Qatar | High income | 65+ threshold | 0.54 | 0.83 (0.78,0.87) | 0.72 (0.62,0.81) | 25 (23,91) | 25 (21,74) |
| Qatar | High income | Full sharing | 0.56 | 0.82 (0.76,0.86) | 0.43 (0.35,0.49) | 24 (21,81) | 17 (13,28) |
| Romania | High income | Default | 0.34 | 0.78 (0.73,0.83) | 0.78 (0.73,0.83) | 297 (169,461) | 297 (169,461) |
| Romania | High income | 2 dose threshold | 0.44 | 0.78 (0.73,0.83) | 0.78 (0.73,0.83) | 297 (169,461) | 297 (169,461) |
| Romania | High income | 40+ threshold | 0.59 | 0.78 (0.72,0.82) | 0.78 (0.72,0.82) | 297 (169,460) | 297 (169,460) |
| Romania | High income | 65+ threshold | 0.58 | 0.78 (0.73,0.82) | 0.73 (0.62,0.81) | 295 (168,455) | 291 (168,448) |
| Romania | High income | Full sharing | 0.53 | 0.79 (0.73,0.84) | 0.38 (0.31,0.47) | 324 (180,511) | 193 (131,251) |
| Russian Federation | Upper middle income | Default | 0.23 | 0.83 (0.74,0.89) | 0.83 (0.74,0.89) | 879 (-1072,2849) | 879 (-1072,2849) |

| Country | Income bracket | Strategy | Proportion vaccinated | Proportion infected | | Mortalities per 100,000 | |
|---|---|---|---|---|---|---|---|
| | | | | unchanged | adapted | unchanged | adapted |
| Russian Federation | Upper middle income | 2 dose threshold | 0.34 | 0.82 (0.72,0.89) | 0.82 (0.72,0.89) | 987 (-724,2848) | 987 (-724,2848) |
| Russian Federation | Upper middle income | 40+ threshold | 0.51 | 0.82 (0.72,0.89) | 0.82 (0.72,0.89) | 976 (-714,2812) | 976 (-715,2813) |
| Russian Federation | Upper middle income | 65+ threshold | 0.53 | 0.81 (0.70,0.89) | 0.70 (0.57,0.81) | 925 (-669,2660) | 838 (-612,2418) |
| Russian Federation | Upper middle income | Full sharing | 0.52 | 0.81 (0.70,0.88) | 0.38 (0.21,0.49) | 925 (-668,2662) | 246 (48,494) |
| Rwanda | Low income | Default | 0.04 | 0.79 (0.57,0.88) | 0.79 (0.57,0.88) | 47 (23,85) | 47 (23,85) |
| Rwanda | Low income | 2 dose threshold | 0.14 | 0.79 (0.57,0.88) | 0.79 (0.57,0.88) | 47 (23,85) | 47 (23,85) |
| Rwanda | Low income | 40+ threshold | 0.30 | 0.79 (0.56,0.88) | 0.79 (0.57,0.88) | 46 (22,84) | 46 (22,84) |
| Rwanda | Low income | 65+ threshold | 0.41 | 0.78 (0.53,0.87) | 0.29 (0.19,0.39) | 43 (20,82) | 25 (14,44) |
| Rwanda | Low income | Full sharing | 0.42 | 0.73 (0.44,0.85) | 0.07 (0.05,0.10) | 34 (17,64) | 7 (5,9) |
| Saudi Arabia | High income | Default | 0.71 | 0.61 (0.45,0.69) | 0.61 (0.45,0.69) | 899 (111,733) | 899 (111,733) |
| Saudi Arabia | High income | 2 dose threshold | 0.81 | 0.61 (0.44,0.69) | 0.61 (0.44,0.69) | 898 (111,732) | 898 (111,732) |
| Saudi Arabia | High income | 40+ threshold | 0.63 | 0.61 (0.44,0.69) | 0.61 (0.44,0.69) | 842 (106,691) | 842 (106,691) |
| Saudi Arabia | High income | 65+ threshold | 0.47 | 0.56 (0.38,0.65) | 0.44 (0.31,0.60) | 597 (79,508) | 506 (70,447) |
| Saudi Arabia | High income | Full sharing | 0.50 | 0.51 (0.34,0.62) | 0.23 (0.18,0.28) | 439 (69,382) | 87 (42,96) |
| Sudan | Low income | Default | 0.02 | 0.65 (0.50,0.74) | 0.65 (0.50,0.74) | 96 (46,198) | 96 (46,198) |
| Sudan | Low income | 2 dose threshold | 0.12 | 0.65 (0.50,0.74) | 0.65 (0.50,0.74) | 96 (46,198) | 96 (46,198) |
| Sudan | Low income | 40+ threshold | 0.28 | 0.64 (0.49,0.74) | 0.64 (0.49,0.74) | 93 (44,191) | 93 (44,192) |
| Sudan | Low income | 65+ threshold | 0.40 | 0.61 (0.43,0.73) | 0.58 (0.38,0.73) | 80 (38,154) | 78 (38,143) |
| Sudan | Low income | Full sharing | 0.42 | 0.57 (0.39,0.72) | 0.32 (0.20,0.43) | 66 (34,121) | 43 (28,59) |
| Senegal | Lower middle income | Default | 0.04 | 0.84 (0.77,0.89) | 0.84 (0.77,0.89) | 75 (48,107) | 75 (48,107) |
| Senegal | Lower middle income | 2 dose threshold | 0.14 | 0.84 (0.77,0.89) | 0.84 (0.77,0.89) | 75 (48,107) | 75 (48,107) |
| Senegal | Lower middle income | 40+ threshold | 0.29 | 0.83 (0.76,0.88) | 0.83 (0.76,0.88) | 74 (47,105) | 74 (47,105) |
| Senegal | Lower middle income | 65+ threshold | 0.39 | 0.78 (0.72,0.84) | 0.62 (0.55,0.68) | 70 (44,100) | 67 (42,97) |
| Senegal | Lower middle income | Full sharing | 0.41 | 0.75 (0.69,0.80) | 0.24 (0.21,0.28) | 61 (39,87) | 32 (23,41) |
| Singapore | High income | Default | 0.89 | 0.49 (0.24,0.66) | 0.49 (0.24,0.66) | 1 (2,4) | 1 (2,4) |
| Singapore | High income | 2 dose threshold | 0.93 | 0.47 (0.15,0.65) | 0.47 (0.15,0.65) | 3 (2,4) | 3 (2,4) |
| Singapore | High income | 40+ threshold | 0.72 | 0.53 (0.20,0.69) | 0.09 (0.07,0.14) | 3 (2,5) | 2 (1,3) |
| Singapore | High income | 65+ threshold | 0.57 | 0.73 (0.57,0.81) | 0.08 (0.06,0.12) | 5 (3,10) | 2 (1,3) |
| Singapore | High income | Full sharing | 0.55 | 0.74 (0.59,0.82) | 0.05 (0.04,0.07) | 5 (3,11) | 1 (1,2) |
| Sierra Leone | Low income | Default | 0.02 | 0.54 (0.39,0.63) | 0.54 (0.39,0.63) | 53 (34,83) | 53 (34,83) |
| Sierra Leone | Low income | 2 dose threshold | 0.12 | 0.54 (0.39,0.63) | 0.54 (0.39,0.63) | 53 (34,83) | 53 (34,83) |
| Sierra Leone | Low income | 40+ threshold | 0.28 | 0.53 (0.38,0.62) | 0.53 (0.38,0.62) | 50 (31,78) | 50 (31,78) |
| Sierra Leone | Low income | 65+ threshold | 0.40 | 0.46 (0.32,0.56) | 0.33 (0.25,0.43) | 39 (25,55) | 36 (25,51) |
| Sierra Leone | Low income | Full sharing | 0.42 | 0.39 (0.27,0.49) | 0.18 (0.14,0.23) | 34 (23,47) | 22 (16,27) |
| El Salvador | Lower middle income | Default | 0.40 | 0.66 (0.52,0.73) | 0.66 (0.52,0.73) | 303 (208,501) | 303 (208,501) |

| Country | Income bracket | Strategy | Proportion vaccinated | Proportion infected | | Mortalities per 100,000 | |
|---|---|---|---|---|---|---|---|
| | | | | unchanged | adapted | unchanged | adapted |
| El Salvador | Lower middle income | 2 dose threshold | 0.48 | 0.66 (0.52,0.73) | 0.66 (0.52,0.73) | 303 (208,500) | 303 (208,500) |
| El Salvador | Lower middle income | 40+ threshold | 0.49 | 0.65 (0.51,0.73) | 0.66 (0.51,0.73) | 299 (206,491) | 299 (206,492) |
| El Salvador | Lower middle income | 65+ threshold | 0.48 | 0.64 (0.50,0.72) | 0.46 (0.32,0.56) | 269 (192,417) | 242 (172,370) |
| El Salvador | Lower middle income | Full sharing | 0.49 | 0.61 (0.45,0.69) | 0.17 (0.15,0.20) | 231 (165,354) | 83 (61,117) |
| Serbia | Upper middle income | Default | 0.45 | 0.77 (0.72,0.80) | 0.77 (0.72,0.80) | 194 (118,303) | 194 (118,303) |
| Serbia | Upper middle income | 2 dose threshold | 0.53 | 0.77 (0.72,0.80) | 0.77 (0.72,0.80) | 194 (118,303) | 194 (118,303) |
| Serbia | Upper middle income | 40+ threshold | 0.65 | 0.76 (0.71,0.80) | 0.76 (0.71,0.80) | 194 (118,302) | 194 (118,302) |
| Serbia | Upper middle income | 65+ threshold | 0.59 | 0.77 (0.73,0.81) | 0.65 (0.56,0.73) | 196 (120,309) | 186 (115,286) |
| Serbia | Upper middle income | Full sharing | 0.53 | 0.79 (0.74,0.82) | 0.31 (0.25,0.35) | 225 (133,361) | 110 (80,137) |
| South Sudan | Low income | Default | 0.01 | 0.49 (0.37,0.62) | 0.49 (0.37,0.62) | 61 (33,94) | 61 (33,94) |
| South Sudan | Low income | 2 dose threshold | 0.11 | 0.49 (0.37,0.62) | 0.49 (0.37,0.62) | 61 (32,94) | 61 (32,94) |
| South Sudan | Low income | 40+ threshold | 0.26 | 0.49 (0.37,0.62) | 0.49 (0.37,0.62) | 60 (32,93) | 60 (32,93) |
| South Sudan | Low income | 65+ threshold | 0.39 | 0.49 (0.36,0.61) | 0.49 (0.36,0.61) | 59 (32,91) | 59 (32,91) |
| South Sudan | Low income | Full sharing | 0.41 | 0.48 (0.35,0.61) | 0.16 (0.11,0.22) | 52 (28,80) | 22 (15,28) |
| Sao Tome and Principe | Lower middle income | Default | 0.15 | 0.73 (0.68,0.77) | 0.73 (0.68,0.77) | 80 (45,136) | 80 (45,136) |
| Sao Tome and Principe | Lower middle income | 2 dose threshold | 0.24 | 0.73 (0.67,0.77) | 0.73 (0.67,0.77) | 80 (45,136) | 80 (45,136) |
| Sao Tome and Principe | Lower middle income | 40+ threshold | 0.32 | 0.72 (0.66,0.76) | 0.71 (0.66,0.76) | 78 (44,134) | 78 (44,134) |
| Sao Tome and Principe | Lower middle income | 65+ threshold | 0.40 | 0.70 (0.63,0.75) | 0.66 (0.58,0.72) | 74 (42,127) | 73 (42,125) |
| Sao Tome and Principe | Lower middle income | Full sharing | 0.41 | 0.68 (0.62,0.74) | 0.44 (0.37,0.51) | 63 (37,105) | 39 (26,60) |
| Suriname | Upper middle income | Default | 0.41 | 0.79 (0.68,0.85) | 0.79 (0.68,0.85) | 233 (110,456) | 233 (110,456) |
| Suriname | Upper middle income | 2 dose threshold | 0.49 | 0.79 (0.68,0.85) | 0.79 (0.68,0.85) | 233 (110,456) | 233 (110,456) |
| Suriname | Upper middle income | 40+ threshold | 0.48 | 0.78 (0.67,0.84) | 0.78 (0.67,0.84) | 228 (107,447) | 228 (107,447) |
| Suriname | Upper middle income | 65+ threshold | 0.48 | 0.77 (0.66,0.83) | 0.71 (0.41,0.84) | 196 (96,378) | 191 (88,384) |
| Suriname | Upper middle income | Full sharing | 0.49 | 0.76 (0.65,0.82) | 0.27 (0.21,0.33) | 160 (79,295) | 70 (38,128) |
| Slovakia | High income | Default | 0.55 | 0.54 (0.44,0.61) | 0.54 (0.44,0.61) | -2506 (-5721,2872) | -2506 (-5721,2872) |
| Slovakia | High income | 2 dose threshold | 0.62 | 0.50 (0.37,0.58) | 0.50 (0.37,0.58) | -960 (-2887,2866) | -960 (-2887,2866) |
| Slovakia | High income | 40+ threshold | 0.67 | 0.49 (0.37,0.58) | 0.49 (0.37,0.58) | -955 (-2874,2853) | -957 (-2878,2858) |
| Slovakia | High income | 65+ threshold | 0.57 | 0.51 (0.38,0.58) | 0.41 (0.30,0.53) | -981 (-2939,2928) | -891 (-2621,2666) |
| Slovakia | High income | Full sharing | 0.53 | 0.52 (0.39,0.60) | 0.13 (0.11,0.15) | -1209 (-3657,3579) | -28 (-140,261) |
| Slovenia | High income | Default | 0.58 | 0.69 (0.61,0.75) | 0.69 (0.61,0.75) | 388 (229,637) | 388 (229,637) |
| Slovenia | High income | 2 dose threshold | 0.65 | 0.69 (0.61,0.75) | 0.69 (0.61,0.75) | 388 (229,637) | 388 (229,637) |
| Slovenia | High income | 40+ threshold | 0.71 | 0.69 (0.61,0.75) | 0.69 (0.61,0.75) | 387 (229,635) | 387 (229,635) |
| Slovenia | High income | 65+ threshold | 0.58 | 0.70 (0.61,0.75) | 0.64 (0.53,0.74) | 385 (228,631) | 377 (225,613) |
| Slovenia | High income | Full sharing | 0.53 | 0.71 (0.62,0.76) | 0.37 (0.25,0.46) | 435 (245,737) | 232 (177,299) |
| Sweden | High income | Default | 0.77 | 0.59 (0.43,0.68) | 0.59 (0.43,0.68) | 488 (263,921) | 488 (263,921) |

| Country | Income bracket | Strategy | Proportion vaccinated | Proportion infected | | Mortalities per 100,000 | |
|---|---|---|---|---|---|---|---|
| | | | | unchanged | adapted | unchanged | adapted |
| Sweden | High income | 2 dose threshold | 0.81 | 0.59 (0.43,0.68) | 0.59 (0.43,0.68) | 488 (263,921) | 488 (263,921) |
| Sweden | High income | 40+ threshold | 0.70 | 0.59 (0.43,0.68) | 0.59 (0.43,0.68) | 486 (262,918) | 487 (263,919) |
| Sweden | High income | 65+ threshold | 0.58 | 0.59 (0.43,0.68) | 0.54 (0.36,0.68) | 475 (261,904) | 463 (252,865) |
| Sweden | High income | Full sharing | 0.52 | 0.61 (0.45,0.70) | 0.19 (0.15,0.23) | 588 (306,1124) | 177 (130,252) |
| Eswatini | Lower middle income | Default | 0.04 | 0.89 (0.85,0.93) | 0.89 (0.85,0.93) | 288 (184,401) | 288 (184,401) |
| Eswatini | Lower middle income | 2 dose threshold | 0.15 | 0.89 (0.85,0.93) | 0.89 (0.85,0.93) | 288 (184,401) | 288 (184,401) |
| Eswatini | Lower middle income | 40+ threshold | 0.31 | 0.88 (0.84,0.92) | 0.88 (0.84,0.92) | 279 (178,389) | 279 (178,389) |
| Eswatini | Lower middle income | 65+ threshold | 0.42 | 0.86 (0.81,0.90) | 0.71 (0.65,0.76) | 259 (164,358) | 246 (153,340) |
| Eswatini | Lower middle income | Full sharing | 0.44 | 0.83 (0.79,0.88) | 0.49 (0.42,0.55) | 224 (144,309) | 137 (96,180) |
| Syria | Low income | Default | 0.01 | 0.51 (0.26,0.68) | 0.51 (0.26,0.68) | 232 (90,547) | 232 (90,547) |
| Syria | Low income | 2 dose threshold | 0.12 | 0.51 (0.25,0.68) | 0.51 (0.25,0.68) | 229 (90,546) | 229 (90,546) |
| Syria | Low income | 40+ threshold | 0.30 | 0.48 (0.21,0.68) | 0.48 (0.21,0.68) | 211 (73,523) | 212 (73,524) |
| Syria | Low income | 65+ threshold | 0.44 | 0.44 (0.16,0.67) | 0.44 (0.15,0.67) | 157 (52,397) | 156 (49,393) |
| Syria | Low income | Full sharing | 0.47 | 0.39 (0.14,0.64) | 0.07 (0.05,0.09) | 97 (38,218) | 24 (16,33) |
| Chad | Low income | Default | 0.00 | 0.48 (0.38,0.57) | 0.48 (0.38,0.57) | 71 (43,109) | 71 (43,109) |
| Chad | Low income | 2 dose threshold | 0.10 | 0.47 (0.38,0.57) | 0.47 (0.38,0.57) | 71 (42,109) | 71 (42,109) |
| Chad | Low income | 40+ threshold | 0.24 | 0.47 (0.37,0.57) | 0.47 (0.37,0.57) | 69 (40,108) | 69 (40,108) |
| Chad | Low income | 65+ threshold | 0.36 | 0.45 (0.36,0.56) | 0.45 (0.34,0.56) | 65 (38,98) | 65 (38,96) |
| Chad | Low income | Full sharing | 0.38 | 0.44 (0.34,0.55) | 0.20 (0.14,0.27) | 55 (33,83) | 26 (19,33) |
| Togo | Low income | Default | 0.05 | 0.61 (0.52,0.68) | 0.61 (0.52,0.68) | 78 (51,140) | 78 (51,140) |
| Togo | Low income | 2 dose threshold | 0.15 | 0.61 (0.52,0.68) | 0.61 (0.52,0.68) | 78 (51,140) | 78 (51,140) |
| Togo | Low income | 40+ threshold | 0.30 | 0.61 (0.51,0.67) | 0.61 (0.51,0.67) | 75 (48,137) | 75 (48,137) |
| Togo | Low income | 65+ threshold | 0.40 | 0.57 (0.41,0.67) | 0.52 (0.31,0.67) | 63 (40,119) | 62 (37,119) |
| Togo | Low income | Full sharing | 0.42 | 0.51 (0.34,0.64) | 0.10 (0.08,0.12) | 47 (31,80) | 15 (12,20) |
| Thailand | Upper middle income | Default | 0.15 | 0.55 (0.36,0.89) | 0.55 (0.36,0.89) | 15 (26,353) | 15 (26,353) |
| Thailand | Upper middle income | 2 dose threshold | 0.27 | 0.55 (0.36,0.89) | 0.55 (0.36,0.89) | 17 (26,353) | 17 (26,353) |
| Thailand | Upper middle income | 40+ threshold | 0.46 | 0.55 (0.35,0.89) | 0.55 (0.35,0.89) | 15 (23,329) | 15 (23,331) |
| Thailand | Upper middle income | 65+ threshold | 0.51 | 0.51 (0.28,0.89) | 0.49 (0.10,0.88) | 4 (12,252) | 4 (7,258) |
| Thailand | Upper middle income | Full sharing | 0.53 | 0.46 (0.20,0.86) | 0.00 (0.00,0.00) | 2 (7,167) | 0 (0,1) |
| Tajikistan | Low income | Default | 0.04 | 0.82 (0.79,0.86) | 0.82 (0.79,0.86) | 93 (59,141) | 93 (59,141) |
| Tajikistan | Low income | 2 dose threshold | 0.15 | 0.82 (0.79,0.86) | 0.82 (0.79,0.86) | 93 (59,141) | 93 (59,141) |
| Tajikistan | Low income | 40+ threshold | 0.29 | 0.82 (0.78,0.86) | 0.82 (0.78,0.86) | 89 (57,135) | 89 (57,136) |
| Tajikistan | Low income | 65+ threshold | 0.41 | 0.80 (0.75,0.85) | 0.77 (0.69,0.84) | 78 (47,121) | 77 (46,121) |
| Tajikistan | Low income | Full sharing | 0.43 | 0.78 (0.73,0.83) | 0.43 (0.37,0.48) | 65 (41,98) | 43 (32,56) |
| Trinidad and Tobago | High income | Default | 0.22 | 0.70 (0.50,0.89) | 0.70 (0.50,0.89) | 307 (104,709) | 307 (104,709) |

| Country | Income bracket | Strategy | Proportion vaccinated | Proportion infected | | Mortalities per 100,000 | |
|---|---|---|---|---|---|---|---|
| | | | | unchanged | adapted | unchanged | adapted |
| Trinidad and Tobago | High income | 2 dose threshold | 0.33 | 0.70 (0.49,0.89) | 0.70 (0.49,0.89) | 307 (104,708) | 307 (104,708) |
| Trinidad and Tobago | High income | 40+ threshold | 0.49 | 0.68 (0.45,0.89) | 0.69 (0.45,0.89) | 297 (101,692) | 298 (101,692) |
| Trinidad and Tobago | High income | 65+ threshold | 0.49 | 0.67 (0.41,0.89) | 0.62 (0.26,0.88) | 253 (86,601) | 249 (79,599) |
| Trinidad and Tobago | High income | Full sharing | 0.52 | 0.63 (0.39,0.86) | 0.09 (0.07,0.14) | 186 (65,441) | 30 (16,53) |
| Tunisia | Lower middle income | Default | 0.16 | 0.87 (0.83,0.90) | 0.87 (0.83,0.90) | 525 (-1443,2002) | 525 (-1443,2002) |
| Tunisia | Lower middle income | 2 dose threshold | 0.27 | 0.87 (0.83,0.90) | 0.87 (0.83,0.90) | 539 (-1361,2001) | 539 (-1361,2001) |
| Tunisia | Lower middle income | 40+ threshold | 0.43 | 0.87 (0.83,0.90) | 0.87 (0.83,0.90) | 526 (-1327,1944) | 527 (-1327,1949) |
| Tunisia | Lower middle income | 65+ threshold | 0.47 | 0.85 (0.81,0.89) | 0.85 (0.79,0.89) | 434 (-1080,1546) | 432 (-1081,1545) |
| Tunisia | Lower middle income | Full sharing | 0.49 | 0.84 (0.78,0.88) | 0.29 (0.24,0.35) | 355 (-791,1202) | 99 (56,157) |
| Turkey | Upper middle income | Default | 0.64 | 0.74 (0.64,0.80) | 0.74 (0.64,0.80) | 136 (53,340) | 136 (53,340) |
| Turkey | Upper middle income | 2 dose threshold | 0.69 | 0.74 (0.64,0.79) | 0.74 (0.64,0.79) | 148 (53,340) | 148 (53,340) |
| Turkey | Upper middle income | 40+ threshold | 0.53 | 0.73 (0.63,0.79) | 0.73 (0.63,0.79) | 147 (53,338) | 147 (53,339) |
| Turkey | Upper middle income | 65+ threshold | 0.51 | 0.75 (0.64,0.80) | 0.63 (0.47,0.74) | 155 (55,371) | 139 (53,306) |
| Turkey | Upper middle income | Full sharing | 0.50 | 0.76 (0.64,0.82) | 0.32 (0.24,0.42) | 157 (57,389) | 64 (43,95) |
| Uganda | Low income | Default | 0.02 | 0.89 (0.68,0.98) | 0.89 (0.68,0.98) | 74 (39,124) | 74 (39,124) |
| Uganda | Low income | 2 dose threshold | 0.12 | 0.89 (0.68,0.98) | 0.89 (0.68,0.98) | 74 (39,124) | 74 (39,124) |
| Uganda | Low income | 40+ threshold | 0.26 | 0.89 (0.68,0.98) | 0.89 (0.68,0.98) | 70 (37,117) | 70 (37,117) |
| Uganda | Low income | 65+ threshold | 0.37 | 0.86 (0.61,0.97) | 0.35 (0.24,0.50) | 44 (25,71) | 26 (16,37) |
| Uganda | Low income | Full sharing | 0.39 | 0.77 (0.47,0.94) | 0.14 (0.12,0.18) | 32 (19,49) | 13 (10,18) |
| Ukraine | Lower middle income | Default | 0.06 | 0.79 (0.69,0.85) | 0.79 (0.69,0.85) | -9 (-1701,1078) | -9 (-1701,1078) |
| Ukraine | Lower middle income | 2 dose threshold | 0.19 | 0.79 (0.69,0.85) | 0.79 (0.69,0.85) | -1 (-1690,1078) | -1 (-1690,1078) |
| Ukraine | Lower middle income | 40+ threshold | 0.38 | 0.79 (0.69,0.85) | 0.79 (0.69,0.85) | -1 (-1679,1071) | -1 (-1679,1071) |
| Ukraine | Lower middle income | 65+ threshold | 0.51 | 0.78 (0.67,0.85) | 0.78 (0.66,0.85) | 6 (-1411,1007) | 5 (-1391,1002) |
| Ukraine | Lower middle income | Full sharing | 0.53 | 0.77 (0.66,0.84) | 0.31 (0.22,0.38) | 23 (-1109,842) | 75 (-2,156) |
| Uruguay | High income | Default | 0.99 | 0.71 (0.53,0.90) | 0.71 (0.53,0.90) | 153 (59,339) | 153 (59,339) |
| Uruguay | High income | 2 dose threshold | 0.90 | 0.71 (0.53,0.90) | 0.71 (0.53,0.90) | 153 (59,338) | 153 (59,338) |
| Uruguay | High income | 40+ threshold | 0.64 | 0.71 (0.53,0.90) | 0.71 (0.53,0.90) | 152 (59,335) | 152 (59,336) |
| Uruguay | High income | 65+ threshold | 0.55 | 0.71 (0.56,0.90) | 0.70 (0.53,0.89) | 143 (55,312) | 141 (55,311) |
| Uruguay | High income | Full sharing | 0.51 | 0.71 (0.58,0.88) | 0.05 (0.04,0.07) | 116 (51,233) | 10 (7,13) |
| United States of America | High income | Default | 0.81 | 0.54 (0.49,0.58) | 0.54 (0.49,0.58) | 300 (244,349) | 300 (244,349) |
| United States of America | High income | 2 dose threshold | 0.84 | 0.54 (0.49,0.58) | 0.54 (0.49,0.58) | 300 (244,349) | 300 (244,349) |
| United States of America | High income | 40+ threshold | 0.68 | 0.68 (0.62,0.73) | 0.47 (0.41,0.54) | 316 (262,364) | 294 (236,347) |
| United States of America | High income | 65+ threshold | 0.57 | 0.79 (0.75,0.82) | 0.46 (0.41,0.53) | 341 (279,393) | 293 (235,345) |
| United States of America | High income | Full sharing | 0.52 | 0.80 (0.77,0.84) | 0.21 (0.20,0.22) | 423 (339,496) | 151 (133,167) |
| Uzbekistan | Lower middle income | Default | 0.11 | 0.79 (0.73,0.83) | 0.79 (0.73,0.83) | 201 (104,471) | 201 (104,471) |

| Country | Income bracket | Strategy | Proportion vaccinated | Proportion infected | | Mortalities per 100,000 | |
|---|---|---|---|---|---|---|---|
| | | | | unchanged | adapted | unchanged | adapted |
| Uzbekistan | Lower middle income | 2 dose threshold | 0.22 | 0.79 (0.73,0.83) | 0.79 (0.73,0.83) | 201 (104,470) | 201 (104,470) |
| Uzbekistan | Lower middle income | 40+ threshold | 0.37 | 0.79 (0.73,0.83) | 0.79 (0.72,0.83) | 195 (101,450) | 195 (101,451) |
| Uzbekistan | Lower middle income | 65+ threshold | 0.45 | 0.77 (0.71,0.82) | 0.73 (0.58,0.82) | 165 (88,342) | 159 (87,305) |
| Uzbekistan | Lower middle income | Full sharing | 0.48 | 0.76 (0.67,0.81) | 0.43 (0.29,0.54) | 138 (76,272) | 87 (62,122) |
| Venezuela | Upper middle income | Default | 0.10 | 0.81 (0.77,0.84) | 0.81 (0.77,0.84) | 290 (232,364) | 290 (232,364) |
| Venezuela | Upper middle income | 2 dose threshold | 0.21 | 0.81 (0.77,0.84) | 0.81 (0.77,0.84) | 290 (232,364) | 290 (232,364) |
| Venezuela | Upper middle income | 40+ threshold | 0.38 | 0.81 (0.77,0.84) | 0.81 (0.77,0.84) | 283 (227,356) | 283 (227,356) |
| Venezuela | Upper middle income | 65+ threshold | 0.46 | 0.78 (0.74,0.82) | 0.70 (0.58,0.78) | 249 (200,313) | 242 (194,310) |
| Venezuela | Upper middle income | Full sharing | 0.48 | 0.76 (0.70,0.81) | 0.33 (0.29,0.37) | 223 (179,277) | 143 (119,172) |
| Vietnam | Lower middle income | Default | 0.05 | 0.46 (0.14,0.71) | 0.46 (0.14,0.71) | 167 (49,307) | 167 (49,307) |
| Vietnam | Lower middle income | 2 dose threshold | 0.17 | 0.46 (0.13,0.71) | 0.46 (0.13,0.71) | 166 (48,307) | 166 (48,307) |
| Vietnam | Lower middle income | 40+ threshold | 0.36 | 0.46 (0.13,0.71) | 0.46 (0.13,0.71) | 147 (37,276) | 148 (37,277) |
| Vietnam | Lower middle income | 65+ threshold | 0.48 | 0.41 (0.08,0.67) | 0.31 (0.02,0.64) | 65 (11,124) | 50 (4,116) |
| Vietnam | Lower middle income | Full sharing | 0.50 | 0.34 (0.04,0.63) | 0.00 (0.00,0.00) | 38 (5,78) | 0 (0,0) |
| Yemen | Low income | Default | 0.01 | 0.68 (0.54,0.81) | 0.68 (0.54,0.81) | 287 (-746,1457) | 287 (-746,1457) |
| Yemen | Low income | 2 dose threshold | 0.12 | 0.68 (0.53,0.80) | 0.68 (0.53,0.80) | 286 (-682,1456) | 286 (-682,1456) |
| Yemen | Low income | 40+ threshold | 0.28 | 0.67 (0.51,0.80) | 0.67 (0.51,0.80) | 255 (-547,1419) | 255 (-547,1421) |
| Yemen | Low income | 65+ threshold | 0.40 | 0.62 (0.44,0.80) | 0.61 (0.41,0.80) | 217 (-416,1250) | 217 (-413,1242) |
| Yemen | Low income | Full sharing | 0.43 | 0.59 (0.39,0.78) | 0.22 (0.14,0.30) | 141 (-222,750) | 38 (17,78) |
| South Africa | Upper middle income | Default | 0.14 | 0.85 (0.77,0.90) | 0.85 (0.77,0.90) | 389 (207,636) | 389 (207,636) |
| South Africa | Upper middle income | 2 dose threshold | 0.25 | 0.85 (0.77,0.90) | 0.85 (0.77,0.90) | 389 (207,636) | 389 (207,636) |
| South Africa | Upper middle income | 40+ threshold | 0.39 | 0.85 (0.77,0.89) | 0.85 (0.77,0.89) | 376 (202,608) | 376 (202,608) |
| South Africa | Upper middle income | 65+ threshold | 0.46 | 0.82 (0.75,0.87) | 0.67 (0.59,0.76) | 331 (180,545) | 306 (167,498) |
| South Africa | Upper middle income | Full sharing | 0.48 | 0.81 (0.73,0.86) | 0.42 (0.35,0.48) | 283 (158,457) | 149 (105,194) |
| Zambia | Lower middle income | Default | 0.01 | 0.87 (0.68,0.95) | 0.87 (0.68,0.95) | 148 (62,296) | 148 (62,296) |
| Zambia | Lower middle income | 2 dose threshold | 0.11 | 0.87 (0.68,0.95) | 0.87 (0.68,0.95) | 148 (62,295) | 148 (62,295) |
| Zambia | Lower middle income | 40+ threshold | 0.26 | 0.87 (0.67,0.94) | 0.87 (0.67,0.94) | 133 (56,266) | 133 (56,266) |
| Zambia | Lower middle income | 65+ threshold | 0.38 | 0.82 (0.58,0.92) | 0.36 (0.23,0.49) | 73 (35,134) | 50 (28,83) |
| Zambia | Lower middle income | Full sharing | 0.40 | 0.73 (0.44,0.87) | 0.16 (0.09,0.23) | 56 (30,95) | 21 (15,28) |
| Zimbabwe | Lower middle income | Default | 0.08 | 0.91 (0.85,0.94) | 0.91 (0.85,0.94) | 80 (46,114) | 80 (46,114) |
| Zimbabwe | Lower middle income | 2 dose threshold | 0.17 | 0.91 (0.85,0.94) | 0.91 (0.85,0.94) | 80 (46,114) | 80 (46,114) |
| Zimbabwe | Lower middle income | 40+ threshold | 0.31 | 0.90 (0.85,0.94) | 0.90 (0.85,0.94) | 77 (44,110) | 77 (44,110) |
| Zimbabwe | Lower middle income | 65+ threshold | 0.39 | 0.87 (0.80,0.91) | 0.53 (0.46,0.61) | 65 (38,92) | 56 (33,79) |
| Zimbabwe | Lower middle income | Full sharing | 0.41 | 0.82 (0.72,0.88) | 0.22 (0.19,0.27) | 57 (34,81) | 24 (17,31) |
