## Supplementary information for "The impacts of increased global vaccine sharing on the COVID-19 pandemic; a retrospective modelling study"

#### Methods and assumptions

We developed a mathematical model of SARS-CoV-2 transmission and COVID-19 disease outcomes, applied to 152 different countries. Each of the 152 countries is simulated using largely independent compartmental infection models – the within country models are only coupled by the global evolution of new variants and the sharing of vaccines when local conditions are met. Individuals within the model may be classified as: susceptible (S); exposed (E); infectious and symptomatic (I); infectious and asymptomatic (A); or recovered (R). We used a set of ordinary differential equations to describe the flow of individuals between these compartments. Susceptible individuals are subject to a force of infection proportional to $I + \tau^a A$, where $\tau^a$ is an age-dependent discounting factor used to represent reduced transmission from asymptomatic individuals compared to symptomatic individuals. The exposed class is further subdivided into three separate states, meaning that in a stochastic formulation the distribution of the latent period would become an Erlang distribution, creating more realistic infection timescales.

Age is recognised to play an important role in the dynamics of the SARS-CoV-2 epidemic, strongly correlating with both the outcome following infection and the characteristic social mixing behavior. To account for these age-based heterogeneities each compartment within the model is stratified into 5 year age groups, from 0-4 years (inclusive) up to a final 100+ years of age group. Each group is populated using country level data on age demographics [1]. Transmission is controlled by the strength of social interactions between age-groups through country dependent who-acquired-infection-from-whom mixing matrices as described by Prem et al. [2]). Additionally, to allow for the effects of NPI measures and social caution, epidemiologically relevant contacts within each country are reduced in a time varying manner by a country-specific control factor $\phi(t)$.

The resulting equations for each country may be given as follows:

$$
\begin{aligned}
\frac{\mathrm{d}S^a}{\mathrm{d}t} &= -\lambda^a \beta^a S^a \qquad \text{where } \lambda^a = \sum_b \phi M^{ab}(I^a + \tau^a A^a) \qquad (1)\\
\frac{\mathrm{d}E_1^a}{\mathrm{d}t} &= \lambda^a S^a - \alpha E_1^a\\
\frac{\mathrm{d}E_2^a}{\mathrm{d}t} &= \alpha E_1^a - \alpha E_2^a\\
\frac{\mathrm{d}E_3^a}{\mathrm{d}t} &= \alpha E_2^a - \alpha E_3^a\\
\frac{\mathrm{d}I^a}{\mathrm{d}t} &= d^a \alpha E_3^a - \gamma^a I^a\\
\frac{\mathrm{d}A^a}{\mathrm{d}t} &= (1 - d^a)\alpha E_3^a \gamma^a A^a\\
\frac{\mathrm{d}R^a}{\mathrm{d}t} &= \gamma^a (I^a + A^a)\,.
\end{aligned}
$$

**Model parameters**

| | |
|---|---|
| $S$ | Number of susceptible individuals |
| $E$ | Number of exposed individuals |
| $I$ | Number of infected symptomatic individuals |
| $A$ | Number of infected asymptomatic individuals |
| $R$ | Number of recovered individuals |
| $\lambda$ | Force of infection |
| $\beta$ | Transmission factor |
| $\alpha$ | Exposure time factor |
| $\phi$ | Behaviour induced transmission scaling |
| $M$ | Mixing matrix |
| $\tau$ | Reduction in asymptomatic transmission |
| $d$ | Probability of exhibiting symptoms |
| $\gamma$ | Recovery rate |

The core infection parameters used are assumed to be the same across all countries. These include age dependent variables for: transmission, $\beta$; the probability of exhibiting symptoms, $d$; the recovery rate, $\gamma$; the reduction in asymptomatic transmission, $\tau$. Estimates for these values are fitted from early age-stratified UK case data to match growth rate, reproductive number and age profiles of infection. Country models vary in demographics, informed by WHO estimates [1], mixing patterns, based on contact matrices described by Prem et al. [2], vaccination levels, mitigating control factors, $\phi$, and death rates, $\psi_d$, amongst cases.

The final parameters, for control and death rates, are fitted at a country level using past daily estimates together with levels of uncertainty for infections and deaths proposed by the Institute for Health Metrics and Evaluation (IHME) [3].

There can be high uncertainty for historic infection estimates in many countries, particularly those that lack the resources and infrastructure for comprehensive surveillance and testing. This uncertainty in the amount of previous infection has important implications for any infection assumption on levels of population immunity, as well as for the correlation with variant growth. Due to this, our model estimates are based on 100 simulations; in each we take a random sample for previous infection and death levels in each country independently from the range informed by the high and low IHME estimates. Control parameters for each sample in each country are then fitted on a six day varying basis using maximum likelihood estimates, and death rate parameters as a least squares fit. Results are given providing both means and 95% prediction intervals informed by these estimates.

To incorporate the effects of variants and changing vaccination levels, the model is run in 2-day time steps. In each time step countries are simulated individually by solving the set of ODEs Eq. (1). Between steps we update a transmission scaling parameter due to variants, $\omega$, as well as country specific vaccine parameters, $v_{inf}$, $v_{tra}$, $v_{sym}$ and $v_{sev}$ scaling infection, transmission, probability of infection being symptomatic and probability of hospitalisation/death respectively for each age group. These are applied to the susceptibility, $v_{inf}\delta$, transmission, $\omega v_{tra}\beta$, and symptomatic probability, $v_{symp}d$, parameters.

The final vaccine parameter, $v_{sev}$, is used to scale severe disease outcomes, used in the calculation of numbers of hospitalisations, $H$, and deaths, $D$, between time steps as follows:

$$H(t + l_h) = v_{sev}(t)\eta_h R(t)$$
$$D(t + l_d) = v_{sev}(t)\eta_d^a R(t)\,,$$

where $\eta_h/\eta_d$ denote the country specific probability of hospitalisation/death and $l_h/l_d$ give the time lag to hospitalisation/death respectively.

Each vaccine parameter is calculated as

$$v(t) = \sum_{s=0}^{t} \upsilon(s)w(t-s)\,,$$

using the amount of vaccination available, $w(t)$, at time step $t$ for each country (as determined by the strategy being followed), together with decaying efficacy parameter, $\upsilon$, with profiles shown in Fig 1 (main text).

| Efficacy against: | 2 doses | 1 dose |
|---|---|---|
| Infection | 75% | 60% |
| Transmission | 45% | 45% |
| Symptoms | 83% | 60% |
| Severe disease | 98% | 80% |

**Table S1:** Efficacy assumptions used, before waning.

The variant transmission parameter is updated to a value relative to estimated global variant prevalence at the time point when total infections up to that time step in simulation match total past infection estimates, that is following the trend of the red line in Fig 1(c) (main text).

Alongside the central scenarios where behaviour is assumed to respect past levels, we additionally explore scenarios where we assume there is reactionary behaviour to control infection levels in countries that begin vaccine sharing. For scenarios with *adapted behaviour*, the control parameter, $\phi$, is left consistent with the *unchanged behaviour* for all countries before they reach the sharing threshold. After this threshold is passed for any particular country, a control scale is first determined by dividing the range of values of $\phi$ used in the *unchanged behaviour* scenarios into 20 even steps, then stepwise increasing/decreasing $\phi$ dependent on whether the number of active infections is increasing/decreasing, subject to a five day time lag.

**(a) Income groups**

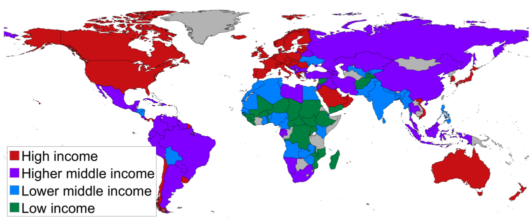


**(b) Vaccination level**

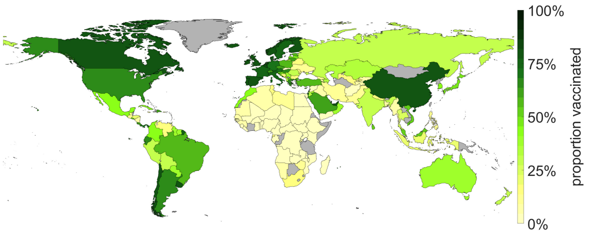


**(c) Infected proportion**

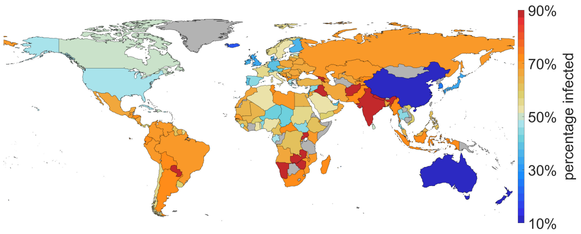


**(d) Deaths per 100,000**

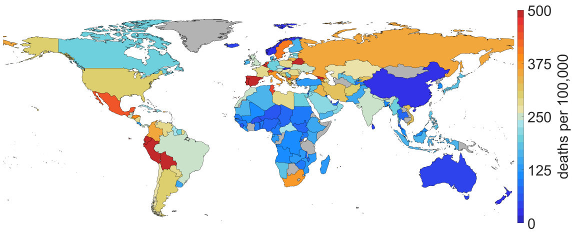


**Figure S1:** Reference maps for the current, low sharing, scenario at the start of 2022. **(a)** income group for each country simulated as defined by the World Bank. Proportion of each simulated country having received full vaccination (2 doses). C estimated proportion of each simulated country to have been infected by SARS-COV-2. Estimated total number of deaths per 100,000 due to COVID-19 disease in each simulated country. In each, grey shading indicates a country that has not been simulated.

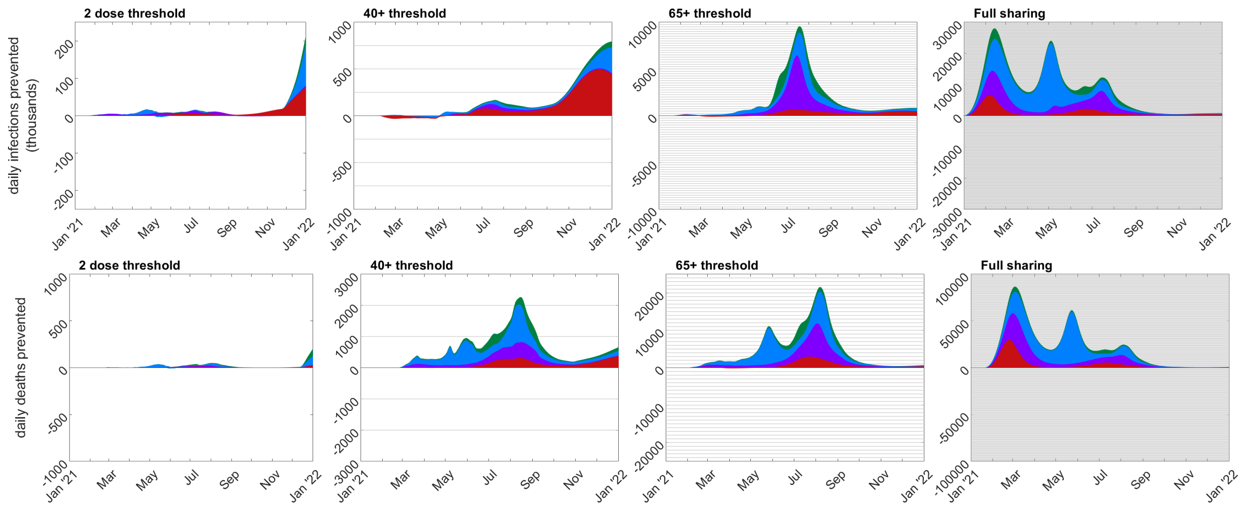


**Figure S2:** Time series plots showing the reduction in the global number of daily infections and daily deaths with adapted behaviour, compared to the default scenario. The different coloured areas represent the proportion of infections/deaths prevented in countries from each economic group: low income (green); lower middle income (blue), higher middle income (purple); high income (red). The columns show the four different vaccine sharing scenarios assuming adapted behaviour.

| **Region** | **Strategy** | **Percentage reduced infection** | | **Percentage reduced mortality** | |
|---|---|---|---|---|---|
| | | unchanged behaviour | adapted behaviour | unchanged behaviour | adapted behaviour |
| World | 2 dose threshold | 0.0 (-0.0,0.3) | 0.1 (0.0,0.4) | -0.3 (-6.3,0.1) | -0.3 (-6.3,0.1) |
| | 40+ threshold | -1.1 (-1.3,-0.6) | 1.1 (0.3,2.4) | 0.6 (-7.3,1.8) | 1.1 (-5.5,2.3) |
| | 65+ threshold | -0.5 (-1.1,1.6) | 12.9 (6.0,19.6) | 7.2 (-6.5,16.4) | 12.3 (-0.9,21.2) |
| | Full sharing | 1.5 (-0.1,4.5) | 64.5 (62.6,65.4) | 11.3 (0.6,23.2) | 62.8 (44.0,76.3) |
| Low income | 2 dose threshold | 0.1 (0.0,0.4) | 0.1 (0.0,0.4) | 0.1 (0.0,0.9) | 0.1 (0.0,0.9) |
| | 40+ threshold | 0.6 (0.2,3.0) | 0.5 (0.2,3.0) | 4.8 (2.8,10.7) | 4.5 (2.8,10.6) |
| | 65+ threshold | 4.6 (2.5,13.4) | 21.7 (12.8,30.3) | 25.3 (16.1,33.1) | 30.2 (20.0,38.2) |
| | Full sharing | 11.5 (5.9,22.1) | 69.1 (65.9,71.4) | 43.5 (34.6,50.2) | 75.1 (64.3,84.8) |
| Lower middle income | 2 dose threshold | 0.0 (0.0,0.2) | 0.0 (0.0,0.2) | 0.0 (-0.9,0.2) | 0.0 (-0.9,0.2) |
| | 40+ threshold | 0.3 (0.1,1.4) | 0.3 (0.1,1.5) | 1.7 (0.7,2.3) | 1.6 (0.5,2.3) |
| | 65+ threshold | 1.7 (0.8,5.2) | 7.1 (3.3,12.2) | 11.4 (9.1,13.1) | 13.5 (11.0,15.6) |
| | Full sharing | 4.0 (1.8,8.2) | 63.6 (62.2,64.9) | 24.5 (21.9,26.4) | 67.4 (62.4,70.6) |
| Higher middle income | 2 dose threshold | 0.0 (0.0,0.5) | 0.0 (0.0,0.5) | 0.0 (-20.6,0.0) | 0.0 (-20.6,0.0) |
| | 40+ threshold | 0.2 (0.1,0.9) | 0.2 (0.1,1.0) | 1.1 (-18.5,2.1) | 1.1 (-18.5,2.1) |
| | 65+ threshold | 1.3 (0.7,4.1) | 19.3 (9.6,29.4) | 7.5 (-16.0,10.6) | 13.6 (-8.9,19.6) |
| | Full sharing | 2.4 (1.3,7.4) | 62.1 (60.4,63.4) | 14.4 (0.3,18.2) | 59.9 (28.7,79.6) |
| High income | 2 dose threshold | 0.0 (-0.1,0.5) | 0.4 (0.1,1.2) | -0.7 (-24.7,0.2) | -0.7 (-24.6,0.2) |
| | 40+ threshold | -10.9 (-18.5,-5.3) | 6.3 (1.1,10.7) | -2.8 (-28.6,2.3) | 0.5 (-22.1,3.3) |
| | 65+ threshold | -17.2 (-31.3,-12.4) | 16.3 (3.5,21.9) | -4.0 (-30.4,24.5) | 5.2 (-18.0,35.4) |
| | Full sharing | -17.2 (-33.4,-13.0) | 68.4 (61.2,71.7) | -23.3 (-62.1,27.7) | 55.4 (26.7,83.9) |

**Table S2:** Estimates for reductions in infection and mortality levels for each vaccine sharing strategy relative to the default scenario. We compute relative estimates for the end of 2021.

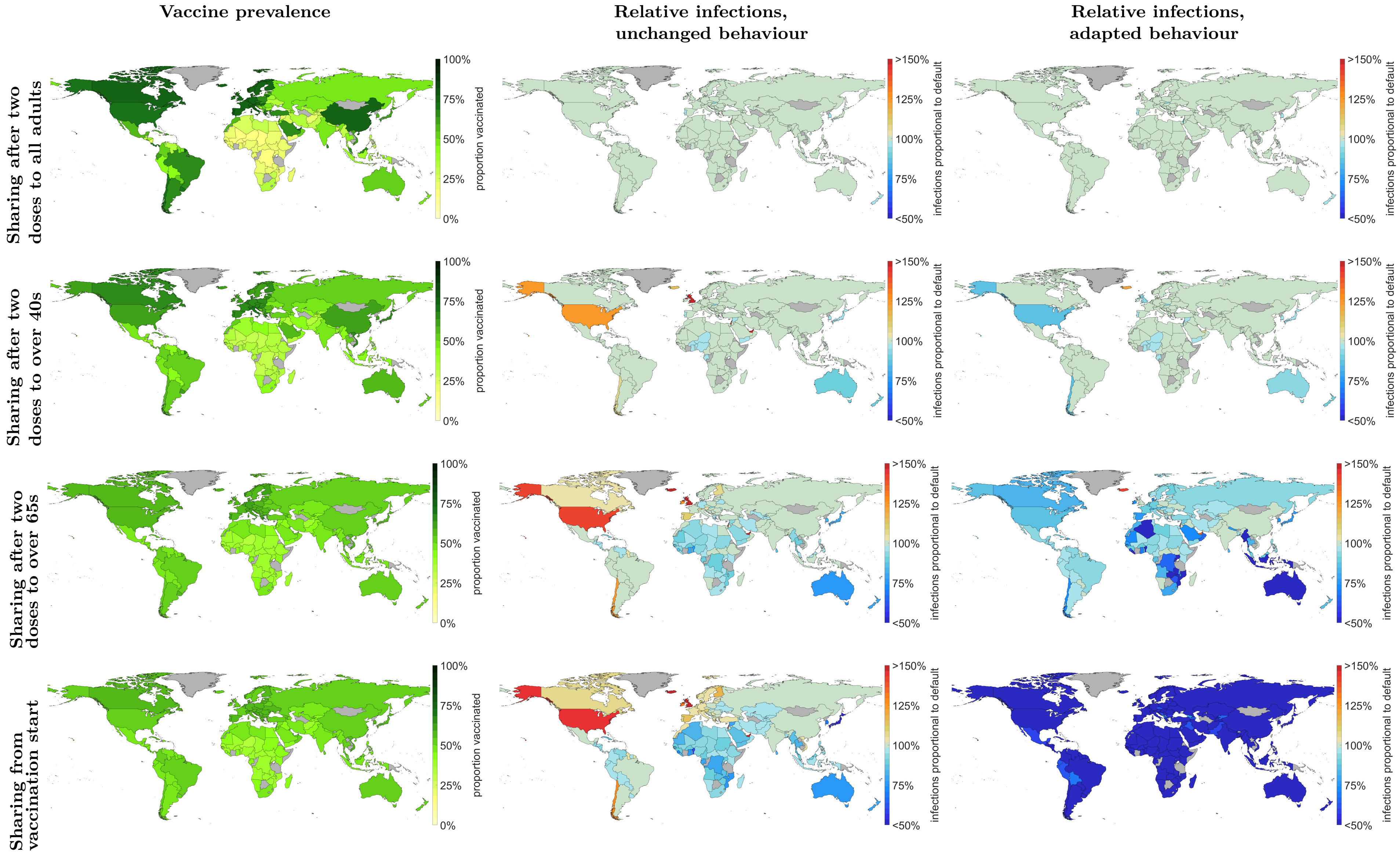


**Figure S3:** Country level estimates of vaccination coverage (left column), total number of infections relative to the current scenario with unchanged behaviour but increased vaccine sharing (central column), and total number of infections relative to the current scenario with unchanged behaviour but increased vaccine sharing (right hand column). All estimates shown for the start of 2022. All values are given for mean simulation values, with 95% prediction intervals provided in Table S2. Any increased degree of vaccine sharing would redress imbalances in global vaccination coverage (left column), resulting in reduced infection and mortality burdens in low, lower middle and higher middle income countries (central and right columns).

### References

[1] World Health Organisation (WHO). Global population demographics, 2021. [Date accessed: 24/01/2022].

[2] Kiesha Prem, Alex R Cook, and Mark Jit. Projecting social contact matrices in 152 countries using contact surveys and demographic data. *PLoS computational biology*, 13(9):e1005697, 2017.

[3] Institute for Health Metrics and Evaluation (IHME). Covid-19 mortality, infection, testing, hospital resource use, and social distancing projections. *University of Washington, Seattle*, 2020. https://covid19.healthdata.org.
